# Cancer incidence and mortality estimates in Arab countries in 2018: A GLOBOCAN data analysis

**DOI:** 10.1101/2022.10.12.22280986

**Authors:** Mariam Al-Muftah, Fares Al-Ejeh

## Abstract

Arab countries are projecting continuing growth in cancer incidence and mortality which require improvements in cancer policies and management. However, there are limited studies that investigate and compare the epidemiology of cancer incidence in Arab countries with only few studies addressing the age-specific rates across cancers. Based on the 2018 estimates from the Global Cancer Observatory, this study provides a comprehensive perspective of cancer burden in 2018 in Arab-speaking countries compared to the rest of the world. The age standardized rates for incidence and mortality for all cancers combined in Arab countries were lower than the global rates but the incidence rates of non-Hodgkin and Hodgkin lymphoma, bladder, breast, and liver cancers were higher. The top-most common cancers and incidence rates, even in age-specific groups, varied between sub-regions of Arab countries (the Levant, Arabian Gulf and Arab African sub-regions), and Iraq and Egypt, suggesting some common and unique environmental factors and possible ethnic or genetic heritages. Arab countries generally had higher mortality-to-incidence ratio than the world’s ratio. This study data is essential reference parameter to evaluate and monitor progress of national initiatives for surveillance programs and clinical management improvements tailored towards reducing cancer incidence and mortality in Arab countries.

## Introduction

An estimated 18.1 million new cancer cases (including non-melanoma skin cancer) and 9.6 million cancer deaths occurred globally in 2018 ^[1, 2]^. It is expected that the global cancer burden will continue to increase to reach 28.4 million cases in 2040 ^[3]^ with an associated rise in social and economic tolls. There are however considerable regional variations in incidence and mortality trends in different regions in the world attributed to the degree of economic development, and social and lifestyle changes ^[1]^. The Arab countries continue to show a rise in cancer incidence with a projected 1.8-fold increase by 2030 ^[4]^. Studies from Saudi Arabia ^[5]^, Egypt ^[6]^, Jordan ^[7]^ and Lebanon ^[8]^ reported an increased age standardized incidence rate (ASIR) during the past 10 years in the five most common cancers. In most Middle Eastern countries (Qatar, Oman, Kuwait, Bahrain, Yemen, Syria, Iraq, Iran, Palestine and Jordan) cancer is the second cause of premature deaths following cardiovascular disease, expect in Lebanon where it is the first cause of death, while in Saudi Arabia and United Arab Emirates, cancer is one of the top 3 causes ^[9]^. In Northern African countries (Algeria, Egypt, Libya, Morocco, Sudan and Tunisia), cancer is ranked as the second cause of death following cardiovascular disease ^[9]^.

Arab countries are contributing to the cancer burden worldwide. For example, in addition to the steady increase in cancer incidence^[8, 10]^, Lebanon has the highest number of bladder cancer cases in the world ^[11]^; the top list also includes Syria and Egypt. Egypt of one of the top contributors to the world’s burden of liver cancer incidence and deaths ^[3]^. Furthermore, increased cancer incidence is expected in the Arab region based on the gradual transition projected between 2000 and 2050 towards an aging population due to reduced fertility and increased life expectancy ^[12, 13]^. Arab countries are classified across different regions in the Global Cancer Observatory (GCO) including Western Asia and Northern, Eastern and Western Africa. While the Middle East and Northern Africa (MENA region) is a geo-political classification and may vary even according to different organizations of the United Nations, it is mainly composed of Arab counties and shares cultural, economic, and environmental similarities. Arab countries in the MENA region may also share ethnic and genetic heritages. Nevertheless, there has been little effort in interrogating cancer trends and incidences in Arab countries in the MENA region. Such a regional/population-based cancer statistics would be instrumental in focusing efforts to decrease cancer burden through systematic implementation of evidence-based interventions for prevention, early diagnosis, and treatment. Such data would be an essential reference parameter to evaluate and monitor progress of national initiatives and surveillance programs tailored towards reducing cancer incidence and mortality.

In this study, we provide a comprehensive analysis of the estimated pattern of incidence and mortality in 2018 of cancers in Arab countries using data from the GCO hosted by the International Agency for Research on Cancer (IARC). There were different trends of cancer incidence and mortality in Arab countries compared to the rest of the World, United States and Europe. Despite differences between Arab countries, we found similarities in cancer incidence rates within sub-regions of the MENA, namely Arab countries of the Levant (Jordan, Palestine, Syria, and Lebanon), Arabian Gulf (Saudi Arabia, Qatar, Kuwait, Oman, Bahrain, and United Arab Emirates but not Iraq) and Africa (Algeria, Libya, Mauritania, Morocco. Somalia, Sudan. and Tunisia but not Egypt). Cancer incidence and mortality rates, even in age-specific groups, were different between the Levant, Arabian Gulf and Arab African subregions and Iraq and Egypt, suggesting some common and unique environmental factors, lifestyle and ethnic or genetic heritages.

## Methods

The global cancer statistics for 2018 from GLOBOCAN 2018 ^[1, 2]^ was interrogated for the estimates of incidence and mortality in Arab countries the Middle East and Northern Africa (MENA) region. Data were accessed through the interactive web-based platform, Global Cancer Observatory (GCO), hosted by the International Agency for Research on Cancer (IARC) ^[14]^. Age-standardized incidence and mortality rates (per 100,000 population), ASIR and ASMR, respectively, are reported in this study and compared between Arab countries, the worldwide (global) rates, and the rates in the United States (US) and Europe. All plots were generated in GraphPad Prism, version 9.4 (GraphPad Software, CA, USA).

## Results

### Overall cancer trends in Arab countries

The age-adjusted rate for incidence (ASIR) of all cancers in 2018 in both sexes at all ages in Arab countries in the MENA region (MENA-Arab) was lower than the global ASIR; 131.9 vs. 197.9 per 100,000 people, respectively (**Figure S1A**). Similarly, the age-adjusted rate for mortality (ASMR) was lower than the global ASMR; 83.5 vs. 101.1 per 100,000 people (**Figure S1A**). The lower ASIR was observed at the cancer site level except for the higher Arab incidence for bladder cancer (8.2 Arab ASIR vs. 5.7 global ASIR), non-Hodgkin’s lymphoma (NHL; 7.2 vs. 5.7), Hodgkin lymphoma (1.7 vs. 0.97), liver cancer (10.6 vs. 9.3) and breast cancer (47.9 vs. 46.3). The mortality-to-incidence ratio (MIR) in 2018 for all cancers in both sexes in most Arab countries was higher compared to the global MIR (**Figure S1B**). It should be noted that MIR is not a proxy of relative cancer survival ^[15]^, however it allows comparisons across countries and regions for mortality rates in relevance to incidence. The top 15 cancers diagnosed in Arab countries were similar to the global trends but differed in their distribution and their contribution to mortality (**Figure S1C**).

Cancer incidence and mortality in Arab countries remained lower than the worldwide rates when separating the data for females and males, but the MIR was higher than the world MIR for most cancer sites (**Figure 1**). Again, the ASIR of NHL and Hodgkin lymphoma, bladder, breast, and liver cancers were higher than the world rates. As shown in **Figure 2**, the incidence of these five cancers varied across the Arab countries, but some sub-regional trends emerged. The higher ASIR of breast cancer was driven by the Levant region (Lebanon, Syria, Jordan, and Palestine). Bladder cancer ASIR was higher than the worldwide ASR in females in Lebanon, Syria, Egypt, and Iraq. In males, while the Levant region, Egypt, Tunisia, Libya, and Algeria had higher ASIR for bladder cancer than the world, the Arabian Gulf countries apart from Iraq (Saudi Arabia, Qatar, Kuwait, Oman, UAE, and Bahrain) had similar or lower ASIR.

**Figure 1:**
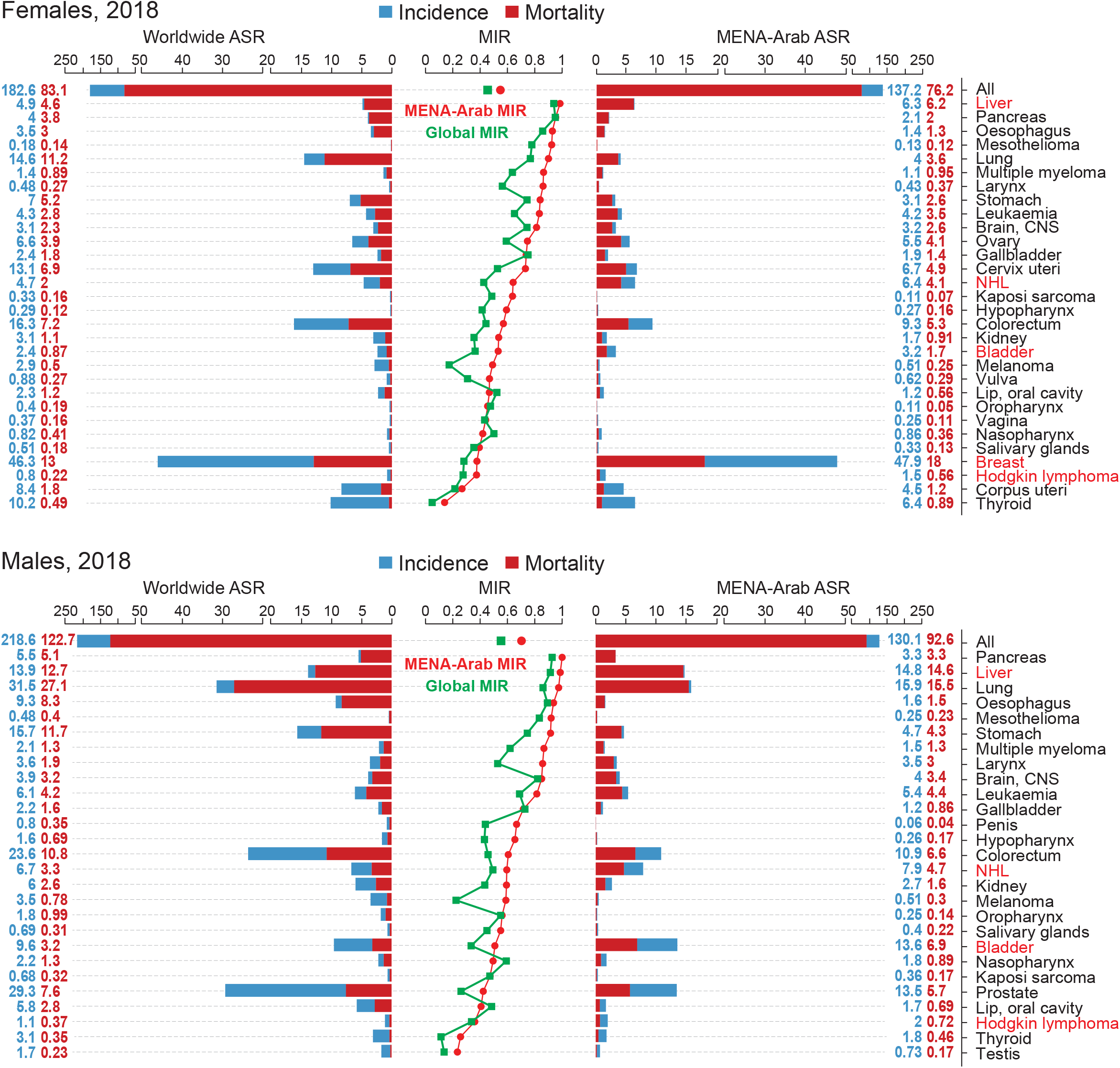
Cancer incidence, mortality, and mortality-to-incidence ratio in the Arab region. The MENA-Arab and worldwide ASR for incidence (blue) and mortality (red) for all cancers and each cancer site for all ages in 2018 for females (top) and males (bottom) are shown. The ASR values for incidence and mortality are shown in blue and red, respectively. Cancer sites are ranked according to the mortality-to-incidence ratio (MIR) shown in the middle line graphs; MENA-Arab MIR in red and worldwide MIR in green. Note: Djibouti, Comoros and Yemen were not included due to missing data.

**Figure 2:**
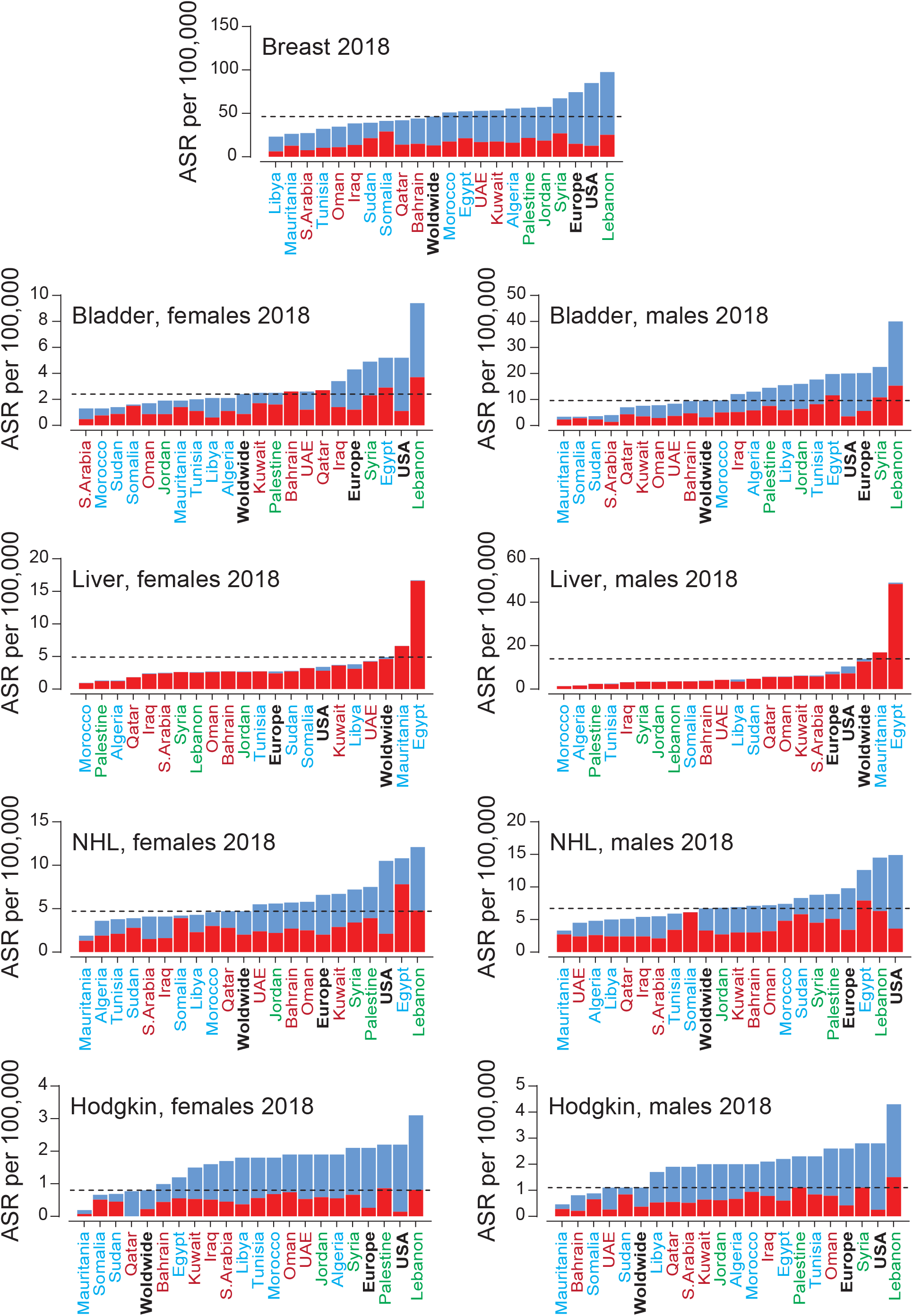
Incidence and mortality rates of five cancers with higher incidence in the Arab region compared to the worldwide incidence. ASR for incidence (blue) and mortality (red) for Arab countries, USA, Europe and worldwide for all ages. The dotted lines mark the worldwide ASR for incidence. Arab countries are labelled as those in the Arabian Gulf (maroon), the Levant (green) and North Africa (blue).

For females and males in Egypt and Mauritania, the incidence of liver cancer was higher than the ASR in the USA, Europe and worldwide (**Figure 2**). For NHL in females and males, Northern African Arab countries except Egypt (Mauritania, Algeria, Tunisia, Sudan, Somalia, Libya, and Morocco) had lower ASIR than the world rate, while the Levant region and half of the Arabian Gulf countries had higher rates. The ASIR of Hodgkin lymphoma in females and males was higher than the worldwide ASIR in most Arab countries. Similar to other cancers, the five cancers with higher ASIR in Arab countries had higher MIR than the worldwide MIR (**Figure S2**), suggesting a scope for better cancer management for several cancer types.

### Leading cancers in females in Arab countries

The top 10 leading cancer sites in females in 2018 for all ages varied across the different Arab countries thus, the top 13 cancers were investigated (**Figure S3A**). The ASIR for all cancers in females was lower than the worldwide ASIR except for Lebanon; ASIR of 248.9 for Lebanon vs. 182.6 worldwide and 134.7 for the rest of the Arab region. Despite some variations, the MIR for the top 13 cancers in females was generally higher in the Arab region compared to the world MIR (**Figure S3B**). As noted earlier, **liver cancer** incidence was markedly higher in Egypt (ASIR 16.7) followed by Mauritania (ASIR 6.6) compared to the rest of the Arab region (ASIR 2.1) and the worldwide rate (ASIR 4.9). The incidence of **lung cancer** was higher in the Levant (ASIR of 9.1) and the world (ASIR 14.6) compared to Arab countries in the Arabian Gulf (ASIR of 3.9) and Northern Africa (ASIR of 3.3). The ASIR for **cervical cancer** in Northern African Arab countries excluding Egypt (ASIR 11.9) was higher than the rest of the Arab region (ASIR 2.7); the worldwide ASIR was 13.10. In contrast, **endometrial cancer** (corpus uteri) was lower in Northern Africa (ASIR 3.5) and Iraq (ASIR 1.8) than the rest of the Arabian Gulf (ASIR 11.8) and the Levant (ASIR 7.5); the world ASIR was 8.40. The incidence was higher for **breast cancer** in the Levant (ASIR 70) compared to the Arabian Gulf (ASIR 35.1), Northern Africa (ASIR 48.4) and the worldwide rate (ASIR 46.3). Similarly, **colorectal cancer** was higher in the Levant region (ASIR 17 vs. 8.5 for the Arabian Gulf and Northern Africa); the world ASIR was 16.30. **Brain cancer** was higher in Iraq (ASIR 4.2), Egypt (ASIR 4.6) and the Levant (ASIR 3.8) compared to the Arabian Gulf (ASIR 1.7), Northern Africa (ASIR 2.3) and the worldwide rate (ASIR 3.1).

### Leading cancers in males in Arab countries

Like in females, the top 10 leading cancer in males for all ages varied across different Arab countries and the top 13 cancers were investigated to represent all countries (**Figure S3C**). Apart from Lebanon (ASIR 240.7), the incidence for all cancers in males in 2018 for the rest of Arab countries (ASIR 125) was lower than the worldwide rate (ASIR 218.6). Like in females, the Arab region had a higher MIR compared to the worldwide MIR (**Figure S3D**). As in females, Egypt had a high incidence of **liver cancer** in males (ASIR 49) compared to the rest of Northern African Arab countries (ASIR 2.8), the Levant (ASIR 3.4), the Arabian Gulf (ASIR 4.7) and the world rate (ASIR 13.9). The incidence of **lung cancer** was lower in the Arabian Gulf (ASIR 9.8) and Northern Africa (ASIR 16.3) than that in the Levant (ASIR of 28.9) and worldwide (ASIR 31.50). Also replicating the trend in females, the incidence of **brain cancer** was higher in males of the Levant (ASIR 5.6), Iraq (ASIR 5.6) and Egypt (ASIR 5.9) compared to the rest of Arab countries in the Arabian Gulf (ASIR 2.3) and Northern Africa (ASIR 3); the worldwide ASIR was 3.90. The incidence of **colorectal cancer** was lower in Iraq (ASIR 7.2), Egypt (ASIR 6.6), Sudan (ASIR 6.2), Mauritania (ASIR 5.2) and Somalia (ASIR 8.4) in comparison to the Levant (ASIR 16.7), the rest of the Arabian Gulf (ASIR 14.1) and Northern Africa (ASIR 13.5), but all were lower than the worldwide rate (ASIR 23.6). The incidence of **bladder cancer** was higher in the Levant (ASIR 25.1), parts of Northern Africa (Egypt, Libya, Algeria, Tunisia, and Morocco: ASIR 15.7) and Iraq (ASIR 12.1) compared to the rest of the Arabian Gulf (ASIR 5) and Northern African countries (ASIR 3.6), and the world rate (ASIR 9.6). **Prostate cancer** was lower in Saudi Arabia (ASIR 6.1), Iraq (ASIR 6.6), Egypt (ASIR 9.5) and Sudan (ASIR 9.2) in comparison to the Levant (ASIR 24.3) and the rest of the Northern African (ASIR 16.7) and the Arabian Gulf (ASIR 15.5) countries and the worldwide rate (ASIR 29.3).

### Sub-regional trends within the Arab countries in females and males

The country-specific analyses above suggest that Arab countries may be divided into subregions; the Levant, Iraq, Egypt, the rest of Arabian Gulf countries (Qatar, Oman, Bahrain, Kuwait, Saudi Arabia, and UAE), and the rest of the Northern African countries (Sudan, Libya, Algeria, Tunisia, Morocco, Mauritania, and Somalia). To this end, these Arab subregions were analysed for the top 10 cancers diagnosed or led to death in 2018 were analysed across females or males at all ages (**Figure S4**). The contribution of cancer types to the total diagnoses varied across the subregions for females and males, suggestion unique genetic, environmental and/or lifestyle differences. Similarly, the contribution of cancer types to cancer-related deaths also varied across the subregions, which may reflect differences in the cancer types diagnosed and/or different patient management in the region.

Next, age-specific ASIR and ASMR for all cancers in 2018 was investigated for all Arab countries in comparison to the worldwide rates, and the rates in the USA and Europe (**Table S1-S2**). Specific age groups were selected for each cancer site to present the trends found for incidence of 30 cancer sites in young, middle, and old age groups (**Figure S5**). These trends are outlined in the following sections.

### Age-specific incidence of blood cancers

Overall, the incidence of blood cancers in many Arab countries was higher than the global rate, particularly at younger age. The incidence of Hodgkin lymphoma in children (under 15 years of age) and young adults (15-34 years) was higher than the worldwide ASIR in most Arab countries for both females and males (**Figure 3A**) and remained higher than the world ASIR in females and males over 35 years of age (**Figure S5**). The ASIR of non-Hodgkin’s lymphoma (NHL, **Figure 3B**) in children, adolescent and young adults was higher in several Arab countries, particularly in females and in Northern African Arab countries. In older Arab females and males, the ASIR for NHL in approximately half of the Arab countries was higher than the world ASIR, and with a trend for higher incidence in Levant countries (**Figure S5**). The incidence of childhood leukaemia in females and males in the Arabian Gulf, Syria and Lebanon was higher than the world ASIR (**Figure 3C**) and remained higher in adults (35 years and older) in the Levant but lower in the Arabian Gulf (**Figure S5**). Multiple myeloma incidence was higher than the world ASIR in 20-54 years old females and males in more than 50% of the Arab countries, particularly in the Levant (**Figure S5**).

**Figure 3:**
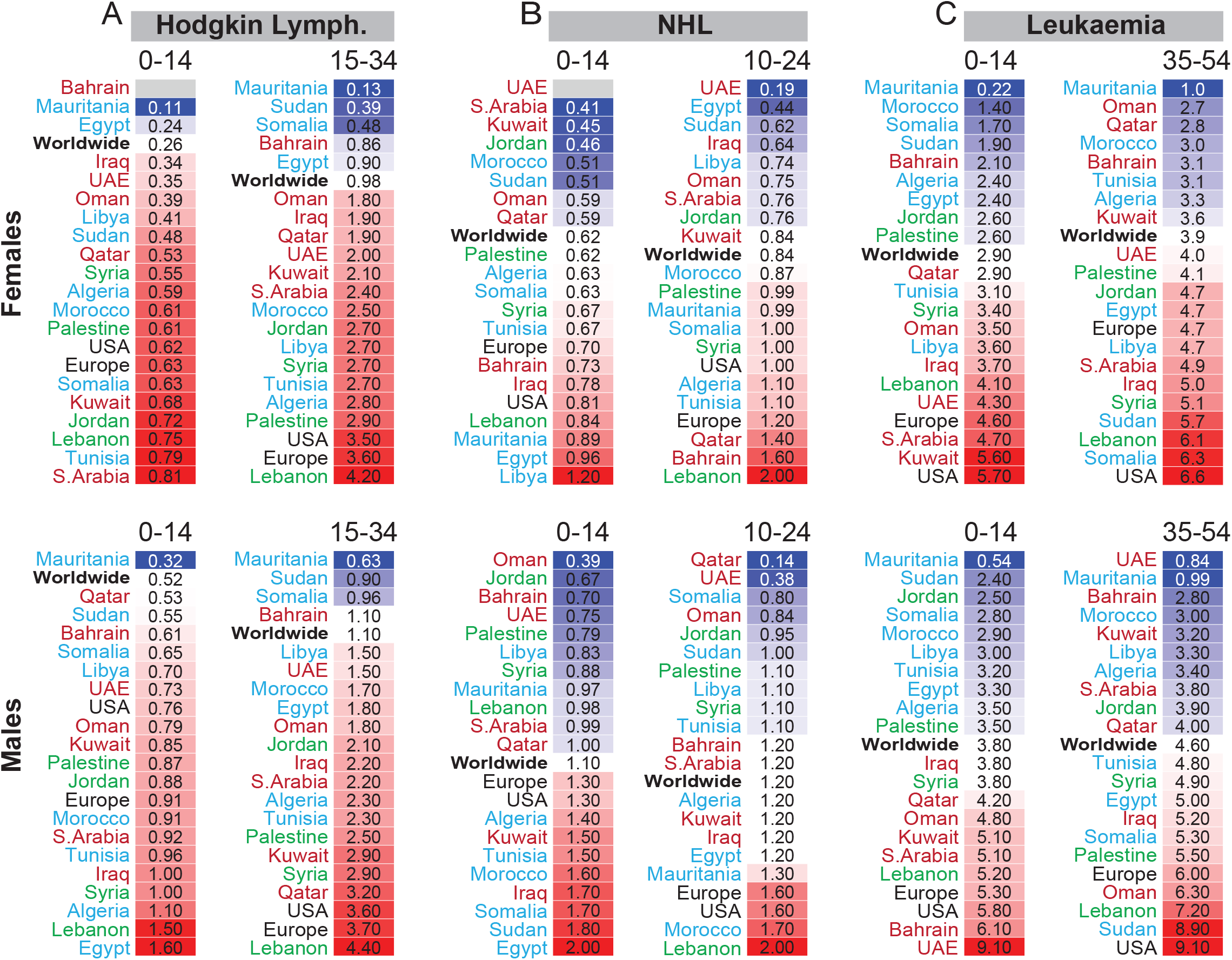
Age-specific incidence of blood cancers in Arab countries. ASIR (**A**) Hodgkin lymphoma, (**B**) non-Hodgkin’s lymphoma (NHL) and (**C**) Leukaemia in females (top panel) and males (bottom panel). Arab countries are labelled as those in the Arabian Gulf (maroon), the Levant (green) and North Africa (blue). The worldwide ASIR is marked with bold font, USA and Europe in black font. ASIR for specific age groups is shown; for all age groups and for multiple myeloma refer to **Figure S5**.

### Age-specific incidence of smoking-related cancers

The incidence of lung cancer (**Figure 4A** and **Figure S5**) in females was lower than the worldwide ASIR in all Arab countries at all ages, except for 20 to 54 years old females in Lebanon. In 20 to 54 years old males, the ASIR of lung cancer was higher than the world rate in Syria, Jordan, Lebanon, Libya, Tunisia, and Morocco. In line with lung cancer incidence in these countries, the incidence of laryngeal cancer (**Figure 4B**) in the 30-54 age group was higher than the world ASIR in the Levant for both females and males, and in Libya, Tunisia, and Morocco for males. When considering bladder cancer (**Figure 4C**), the higher incidence in Syria, Lebanon, Jordan, and Iraq in females between 20 and 54 years old compared to the world ASIR is also in line with the higher incidence of lung and laryngeal cancers in these countries. These trends suggest tobacco-related causes. Similarly, the higher incidence of bladder cancer in 20 to 54 years old males in 12 out of 19 Arab countries compared to the world ASIR may also be explained by tobacco use. However, three countries with high bladder cancer incidence in the 20-54 age group stand out: Mauritania for females, Sudan for males, and Egypt for females and males. The ASIR for lung and laryngeal cancers in those countries were lower than the world ASIR, suggesting other factors behind the higher bladder cancer incidence. Similarly, the higher bladder cancer incidence in 55 to 69 years old females in several Arabian Gulf countries compared to the world ASIR (**Figure 4C**) was at odds with the lower incidence of lung and laryngeal cancers, again suggesting other factors such as exposure to chemicals other than tobacco smoke.

**Figure 4:**
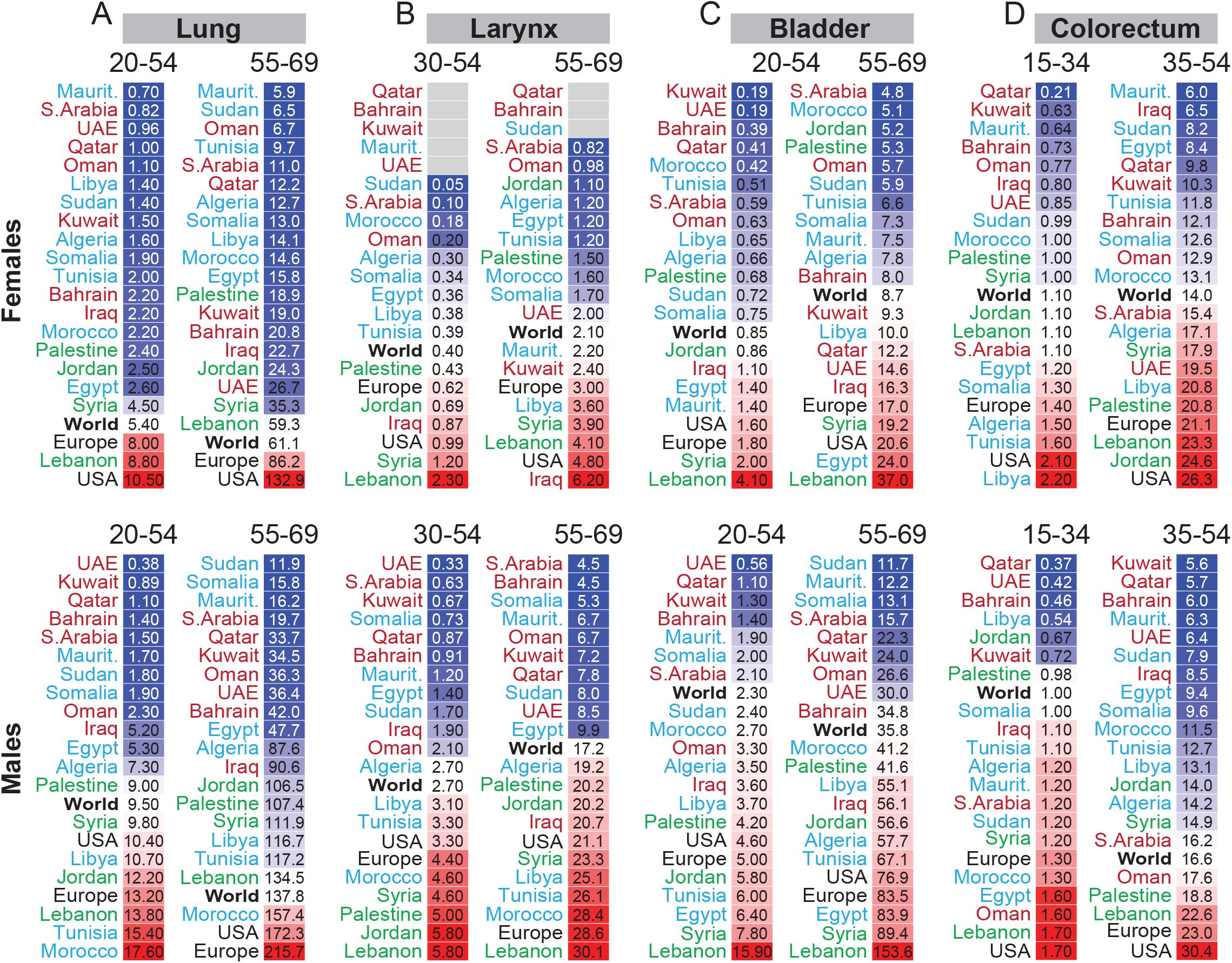
Age-specific incidence of smoking-related cancers in Arab countries. ASIR for cancers of the (**A**) lung, (B) larynx, (**C**) bladder and (**D**) colorectum in females (top panel) and males (bottom panel). Arab countries are labelled as those in the Arabian Gulf (maroon), the Levant (green) and North Africa (blue). The worldwide ASIR is marked with bold font, USA and Europe in black font. ASIR for specific age groups is shown; for all age groups refer to **Figure S5**.

The ASIR for colorectal cancer in 15 to 34 years old females and males (**Figure 4D**) was higher than the world ASIR in several Northern African Arab countries, Saudi Arabia, Oman, and Iraq. The young age and the relatively lower incidence of lung and laryngeal cancers in those countries suggest a non-tobacco related cause; most likely diet since alcohol consumption in Arab countries is low. The incidence of colorectal cancer in older Arab males, particularly over 55 years old (**Figure S5**), was lower than the world ASIR except of 35 to 54 years old males in Oman, Palestine, and Lebanon (**Figure 4D**). For females between 35 and 54 years of age, the ASIR for colorectal cancer in the Levant, Libya, Algeria, UAE, and Saudi Arabia was higher than the world rate. Interestingly, and relative to the world ASIR, the higher incidence of colorectal cancer at young age and the lower incidence at older age (over 55 years, **Figure S5**) in Arabs might reflect a generational change in diet rather than tobacco or alcohol consumption.

### Age-specific incidence of sex-specific cancers

Cervical cancer (**Figure 5A**) had an alarming ASIR in four Northern African Arab countries; Libya, Morocco, Somalia and Mauritania, and the incidence in Morocco, Somalia and Mauritania was higher than the worldwide rate in females between 35 and 54 years of age (**Figure S5**). These trends implicate human papillomavirus (HPV) infections, which is supported by the high ASIR of vaginal cancer in those Northern African countries (**Figure S5**). The high incidence of oropharyngeal, nasopharyngeal, and oesophageal cancers in Northern African Arab countries in both females and males (**Figure S5**), particularly in Sudan, Somalia, Libya, Tunisia, Algeria, and Morocco, further illustrate the impact of HPV prevalence in North Africa.

**Figure 5:**
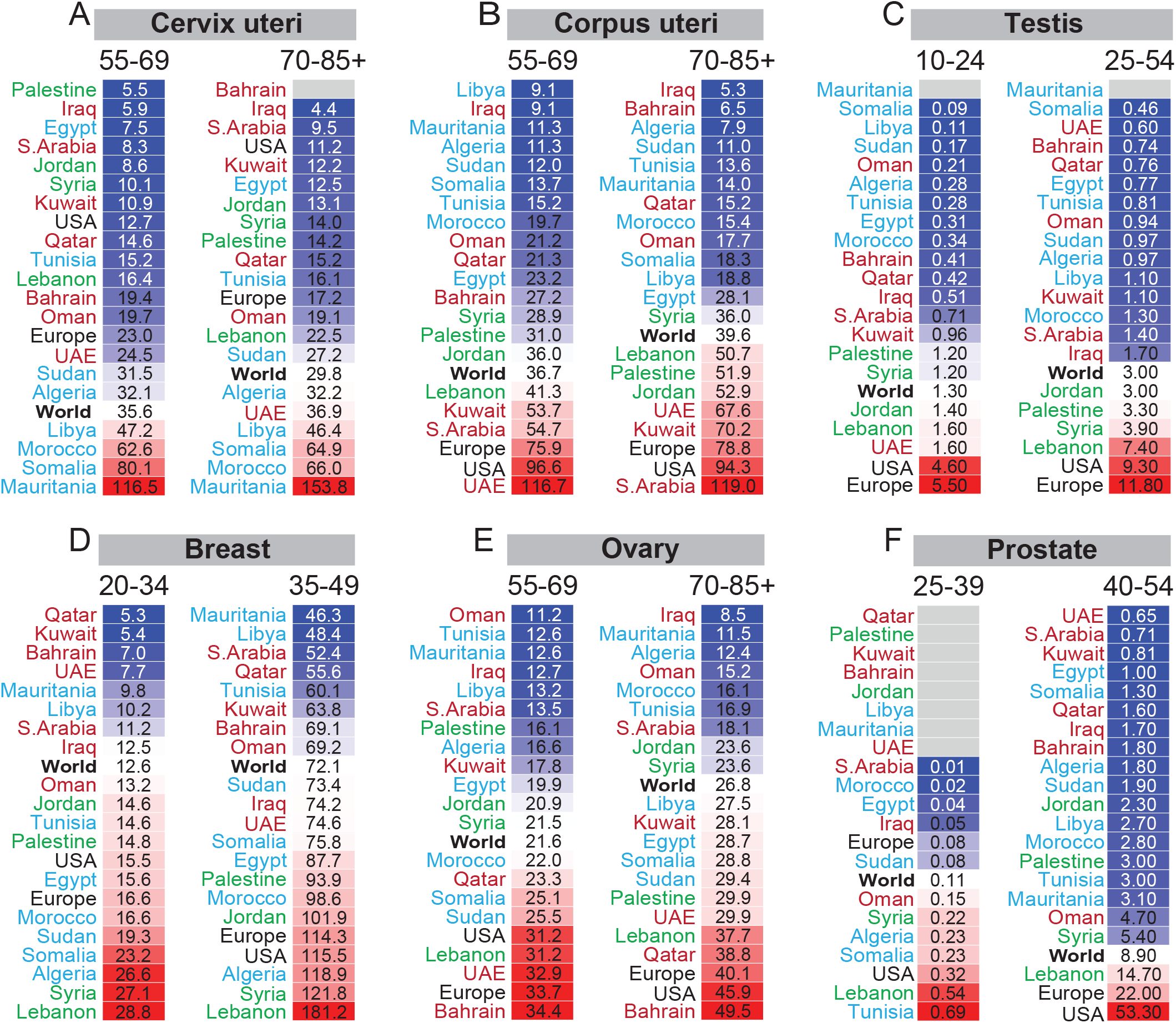
Age-specific incidence of sex-specific cancers in Arab countries. ASIR for sex-specific cancers in females (top panel) and males (bottom panel). Arab countries are labelled as those in the Arabian Gulf (maroon), the Levant (green) and North Africa (blue). The worldwide ASIR is marked with bold font, USA and Europe in black font. ASIR for specific age groups are shown; for all age groups and for other sex-specific cancers refer to **Figure S5**.

The ASIR for endometrial cancer was higher in the Levant and three Arabian Gulf countries (UAE, Saudi Arabia, and Kuwait) than Northern African Arab countries at all ages (**Figure S5**). Endometrial cancer ASIR in the Levant countries, UAE, Saudi Arabia, and Kuwait exceeded the world rate in females above 55 years of age (**Figure 5B**). Testicular cancer ASIR (**Figure 5C**) was generally lower in Arab countries compared to the world rate, but young adults in the Levant countries had higher incidence than the world ASIR.

The ASIR for breast cancer (**Figure 5D**) in females under 50 years of age was higher than the world ASIR in 11 out of 19 Arab countries mainly in the Levant and Northern African Arab countries. The Arabian Gulf countries had lower ASIR for breast cancer in females under 50 years of age. Breast cancer incidence in females aged 50 years and over remained higher than the world ASIR in the Levant and Egypt, and the ASIR in Bahrain, UAE, Kuwait, and Qatar increased over the world rate (**Figure S5**). The incidence of ovarian cancer in females under 55 years was lower in most Arab countries (17 out of 19) than the world ASIR (**Figure S5**). Ovarian cancer incidence increased in older females (55-69 years and over 69 years) in several Arab countries (**Figure 5E**). The ASIR for prostate cancer in the age groups of 55-69 years and over 69 years was higher than the world rate only in Lebanon and Kuwait and Lebanon, respectively (**Figure S5**). Similarly, in men between 40 and 54 years of age, only Lebanon exceeded the world ASIR for prostate cancer while the ASIR for the rest of the Arab countries was considerably lower (**Figure 5F**). In young men, under 40 years, six Arab countries (Oman, Syria, Algeria, Somalia, Lebanon, and Tunisia) had higher incidence of prostate cancer than the world ASIR, and the ASIR in Lebanon and Tunisia exceeded the ASIR in the USA (**Figure 5F**).

The incidence of breast cancer in young Arab women and prostate cancer in young Arab men point to hereditary factors. To gain more insight in addition to the trends for ovarian cancer, we focused on the incidence of pancreatic and stomach cancers and melanoma. The incidence of pancreatic cancer in the 35-49 years age group (**Figure 6A**) was higher than the world ASIR for females in the Levant, Morocco, Sudan, Egypt, Somalia, and Libya, and for males in the Levant, Tunisia, Egypt, and Libya. In contrast, the incidence of pancreatic cancer in 50 years or older females and males in most Arab countries (except females in Kuwait, UAE, and Bahrain) was lower than the world ASIR (**Figure 6A** and **Figure S5**). Like pancreatic cancer, the incidence of gastric cancer in young (20-39 years) females and males was higher than the world, USA, or European ASIR in the Levant, Morocco, Tunisia, Algeria, Bahrain, and Oman (**Figure S5**). The incidence of melanoma in all Arab countries at all ages, except for 20-54 years old males in Lebanon and Palestine, was lower than the world, USA, and European ASIR (**Figure S5**). However, it is noteworthy that the Levant, Tunisia, Algeria, Somalia, Oman, UAE, and Kuwait had higher melanoma ASIR in females and males under 55 years of age compared to the rest of the Arab countries. Altogether, the trends of breast, ovarian, prostate, pancreatic and stomach cancers and melanoma in young females and males in certain Arab countries support *BRCA1*- and *BRCA2*-associated hereditary breast and ovarian cancer syndrome (HBOC).

**Figure 6:**
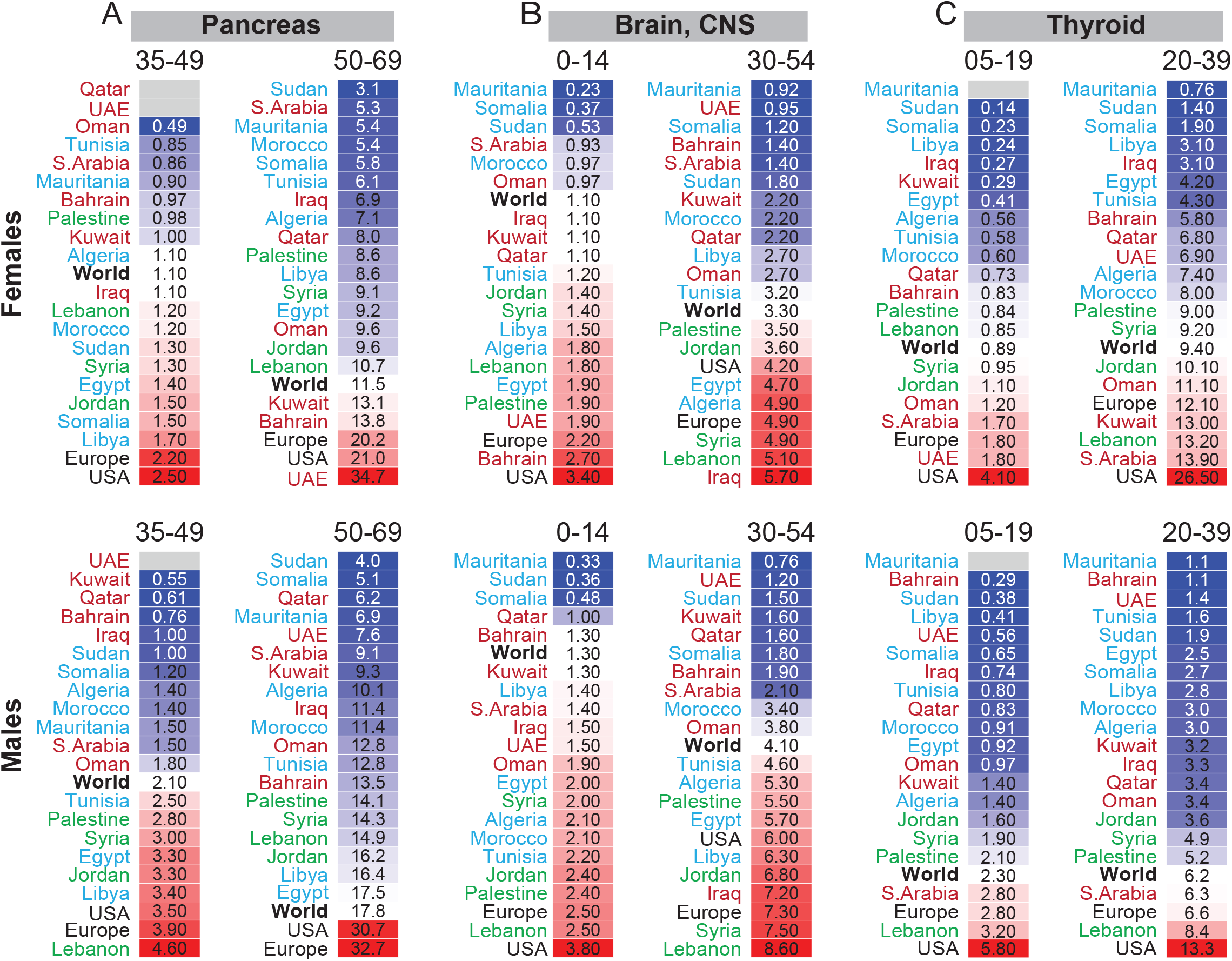
Age-specific incidence of other solid cancers in Arab countries. ASIR for cancers of the (**A**) pancreas, (**B**) Brain, central nervous system (CNS), and (**C**) thyroid n females (top panel) and males (bottom panel). Arab countries are labelled as those in the Arabian Gulf (maroon), the Levant (green) and North Africa (blue). The worldwide ASIR is marked with bold font, USA and Europe in black font. ASIR for specific age groups is shown; for all age groups and other cancer sites refer to **Figure S5**.

### Age-specific incidence of other solid cancers

The incidence of brain cancer (**Figure 6B**) in Arab children (under 15 years) was similar or higher than the world ASIR in 13 and 15 out the 19 Arab countries analysed for females and males, respectively. This higher ASIR continued for young adults (15 to 29 years, **Figure S5**) and 30 to 50 years fold females and males (**Figure 6B**) in the Levant, Iraq, Tunisia, Algeria, and Egypt.

The incidence of thyroid cancer in 5 to 19 and 20 to 39 years old females was higher than the world ASIR in Saudi Arabia, Kuwait, UAE and Oman from the Arabian Gulf, and Syria, Jordan, and Lebanon from the Levant (**Figure 6C** and **Figure S5**). In male children and young male adults (less than 40 years of age), the incidence of thyroid cancer only in Saudi Arabia and Lebanon was higher than the world ASIR (**Figure 6C**). For over 39 years of age (**Figure S5**), the incidence of thyroid cancer was higher than the world ASIR for males in Saudi Arabia, Oman, UAE, Kuwait, Bahrain, Sudan, and Lebanon, and for females in Saudi Arabia, Kuwait, UAE, Lebanon, Jordan, Palestine, Morocco, and Libya.

## Discussion

The 2018 ASIR for incidence and mortality for all cancers combined in both sexes in Arab countries were lower than the global rates. This trend was observed at the organ-specific level except for the higher incidence of Hodgkin and non-Hodgkin’s lymphomas and cancers of the bladder, liver, and breast. The mortality-to-incidence ratio, MIR, in Arab countries was higher, suggesting a scope for improving cancer management in the region.

Considering that cancer is an age-related disease, the generational and demographical structure in Arab countries cannot be ignored as a factor behind the lower cancer incidence. Older Arabs (55 to 75 years and older) diagnosed with cancer in 2018 were born between 1943 and 1963; a period after the second world war when most of these countries were gaining independence. These generations had a different lifestyle and less affected by modernisation and industrialisation seen in other regions in the world such as Europe and the USA. Recently, modern lifestyles in the Arab region have started to impact obesity, including childhood obesity, especially in the Arabian Gulf area ^[16]^. In addition to the link between obesity and some cancers, the transition to western lifestyles in the Arab region may change the profile of cancer incidence the future. Furthermore, the Arab region showed a delayed decline in infertility compared to the rest of the world, and it is projected that by 2050 there will be several folds increase in the proportion of the population aged above 65 years old ^[12]^. This provides a window of opportunity for Arab governments to early place effective policies to tackle such challenge that will impact cancer incidence. The effect of those new, more modern lifestyles in the Arab region may be captured by the next releases of the GLOBOCAN data in comparison to the portrait of cancer trends in Arab countries in 2018 reported here.

Cancer incidence in Arab countries in 2018 revealed clear trends such as the high incidence of HPV-related cancers (cervical, vaginal, oropharyngeal, nasopharyngeal, and oesophageal cancers) in Northern African Arab countries. Similarly, the high incidence of liver cancer in 2018 in Northern African Arab countries, particularly Egypt and Mauritania, can be attributed to the high prevalence of the human oncogenic viruses hepatitis B or C virus (HBV or HCV) ^[17-19]^. The high prevalence of HCV in Northern Africa may also relate to the higher incidence of non-Hodgkin lymphoma in boys and girls under 15 years of age in Egypt and several Northern African Arab countries in 2018. In support, HCV infection has been shown to increase the risk by 14-fold for developing non-Hodgkin’s lymphoma in Egyptian patients ^[20]^. HPV and HBV vaccination and treating hepatitis chronic inflammation to avoid liver scarring (cirrhosis), are obvious approaches to reduce the burden of these virus-related cancers. The safety and efficacy of HPV vaccines against infection by cervical cancer-associated HPV types have been demonstrated by international randomized clinical trials. High income countries with high vaccinations coverage observed substantial decline of HPV infections in girls and young women up to 8 years post vaccination ^[21]^. Modelling analysis predicts that high-coverage girls-only HPV vaccination with once or twice lifetime screening can eliminate cervical cancer as a public health issue in low income and lower-middle-income countries including in North Africa and the Middle East ^[22]^.

Most Arab countries, except Mauritania and Bahrain, showed higher ASIR in 2018 than the worldwide incidence of Hodgkin’s lymphoma in both sexes in children and adults. Although Epstein-Barr virus (EBV) has been suspected to increase the incidence of Hodgkin’s lymphoma in developing countries, studies from Kuwait, Saudi Arabia and UAE reported EBV inflection positivity in patients close and the worldwide incidence and that genetic susceptibility may play a role ^[23-25]^.

The low incidence of lung, laryngeal and bladder cancers in females from most Arab countries is in line with the lower prevalence of tobacco use in females; the maximum age-standardized prevalence estimates of tobacco use in females between 2007 and 2018 was 6% and 3.6%, respectively, in Algeria, Egypt, Iraq, Kuwait, Morocco, Oman, Qatar, Saudi Arabia, Tunisia, and UAE ^[26]^. The high incidence of lung, laryngeal and bladder cancers in Lebanese females may be explained by the considerably higher prevalence of their tobacco use (36.5% in 2007 to 35.9% in 2018) ^[26]^. These three tobacco use-related cancers had high incidence in 2018 in males of the Levant region particularly Lebanon, which is in line with high prevalence tobacco use in Lebanon (49.5% in 2007 to 49.4% in 2018) ^[26]^, and in Jordan (59% in 2006 to 42% in 2009) and Syria (40% in 2006 to 35% in 2009) ^[27]^. From the North African Arab countries, males from Tunisia and Morocco had the highest incidence of lung, laryngeal and bladder cancers and the highest prevalence of tobacco use; Tunisia 62.6% to 49.1% and Morocco 37.4% to 28.6% in 2007 to 2018, respectively ^[26]^.

While several Arab countries showed lower than the world’s ASIR for lung and laryngeal cancers in 2018, they presented with higher ASIR for bladder and/or colorectal cancers suggesting other causes than tobacco-use. Unfortunately, there are insufficient studies investigating the genetics and mutational profiles in Arab countries to understand the epidemiology of bladder cancer and subsequently guide targeted public health and cancer screening programs. Tobacco smoking is one of the most important risk factors contributing to the development of bladder cancer, followed by occupational exposure to aromatic amines, polycyclic aromatic hydrogens, and chlorinated hydrogens. Drinking water contaminated with halogenated chemical species has contributed to 8.6 % of bladder cancer cases in Lebanon ^[28]^. The n increasing trend in colorectal cancer in young Arab adults (below 50 years old) ^[29-31]^ may be driven by the rapid environmental, lifestyle and industrialization changes in the last two decades. The average prevalence of obesity has drastically increased in Arab countries in the past three decades (6.5% in 1975 to 20% in 2016) which has been associated with unique genetic polymorphism in Arabs ^[32]^. This increase in obesity might explain the increase of colorectal cancer in younger Arabs given the association of diabetes with higher risk for colorectal cancer ^[33]^, including younger women ^[34]^.

Most Arab countries showed higher than the world’s ASIR for breast cancer in females under 50 years of age. It has been reported that between 1950-2008 the average age of presentation of breast cancer in 11 Arab countries (Egypt, Jordan, United Arab Emirates, Kuwait, Lebanon, Oman, Qatar, Saudi Arabia, Sudan, Tunisia, and Yemen) was a decade earlier than in Western countries ^[35]^. Genetic hereditary factors may be the cause for this, which is supported by the higher ASIR in Arab countries than the world ASIR at younger age for cancers associated with *BRCA* mutations including ovarian, pancreatic, gastric, and prostate cancers. Several studies on Arabs reported high number of mutations and novel pathogenic mutations in *BRCA1* and *BRCA2* genes in breast cancer patients ^[36-42]^. A Meta-analysis of reported prevalence of *BRCA* mutations in Arab countries demonstrated that one in five hereditary breast and/or ovarian cancer patients are likely to bare *BRCA* mutations, and the Levant region showed higher prevalence of *BRCA* mutations compared to other Arab countries; however, this meta-analysis had high heterogeneity ^[43]^.

Large scale studies to investigate the mutational spectrum of *BRCA1* and *BRCA2* mutations in the general Arab population, and in cancer patients, and identifying founder mutations will have profound implications in patient screening and management to reduce incidence of *BRCA* mutation associated cancers. This is particularly important considering the various ethnicities in Arabic-speaking countries. One example of such a study was based on the Qatar Genome Programme cohort where whole genome sequencing was carried out on more than 6,000 individuals across six ancestry groups in Qatar; general Arabs, Eastern or Persian, admixture of Arab subpopulations, Arabian peninsula, African, and South Asian ^[44]^. This study found that 56.4% of the identified *BRCA1/BRCA2* variant carriers were in Qataris of Persian origin, and those pathogenic variants were completely absent in Qataris of Arabian Peninsula origin ^[44]^. Qataris with Arabian Peninsula ancestry also showed the lowest polygenic risk score mean for colorectal cancer while those of African ancestry had the highest ^[44]^. This study from Qatar points to one limitation of our study here regarding the genetic risk for certain cancers, such as breast, ovarian and colorectal cancers since Arabic-speaking countries span across two continents and have different ethnic compositions across and possibly within these countries. The heterogenous ethnic composition of Arab countries would not influence viral infection-related cancers such as liver and cervical cancers. Another limitation of our study is the variation across the MENA region, including Arab countries, in terms of the human development index (HDI) which are associated with different types of cancers at varying magnitudes ^[1, 2]^.

This study of the estimated cancer incidence and mortality rates in Arab countries for 2018 from the GLOBOCAN provides a baseline for future analyses of the next releases of the GLOBOCAN estimates to follow trends over time. We also shed some light on risk factors that may associate with cancers in the Arab countries, particularly based on the analysis of age-standardized rates across different age groups which emphasized the need for more well-designed genetic epidemiologic studies. In addition to the abovementioned ethnic diversities across and within Arab countries, consanguineous marriages which accounts for 35-50 % of marriages in Arab countries ^[45-47]^ further underlines the importance of genetic epidemiology studies for the region. Finally, based on the mortality to incidence ratio, the study identified a scope for improving cancer treatment in the Arab region generally, and for certain Arab countries. In conclusion, much effort is required to identify the genetic and mutational landscapes of the Arab population to better understand genetic risks for cancer and to guide cancer management for reducing cancer burden in this region.

## Data Availability

All data produced are available in the Supplementary Data file included in the submission.

## Notes

### Competing Interest Statement

The authors have declared no competing interest.

### Funding Statement

This study was funded by Qatar Biomedical Research Institute (QBRI) - Hamad bin Khalifa University (HBKU), Qatar Foundation, Doha, Qatar.

