## Supplementary Figures for "Cancer incidence and mortality estimates in Arab countries in 2018: A GLOBOCAN data analysis"

**Supplementary Figures and Supplementary Data**

**Figure S1**: Age-standardized rate (ASR) for cancer incidence and mortality, and mortality-to-incidence ratio (MIR) in both sexes in 2018.

**Figure S2**: Mortality-to-incidence ratio (MIR) for five cancers with higher incidence in the Arab region.

**Figure S3**: The top cancer sites in the MENA-Arab region leading incidence or mortality in females or males in 2018 for all ages.

**Figure S4**: Leading cancers for diagnosis and cancer-related deaths in subregions of the Arab countries.

**Figure S5**: Age-specific incidence of cancers in Arab countries.

**Supplementary Data**: The extracted GLOBOCAN 2018 used in the study are summarized in **Table S1** which allows filtering for cancer types, sex, type of data (ASR for incidence or mortality), and different age groups across Arab countries, Arab regions, the world, USA and Europe. **Table S2** in this **Supplementary Data** file contains the data behind Table S1 with additional information; the number of cases and crude rates. Note: Tables S1 and S2 are locked to protect the integrity of the data but users can filter the data and copy results from Table S1, and copy the original data from Table S2; no other editing is permitted.

**Supplementary Figure 1**

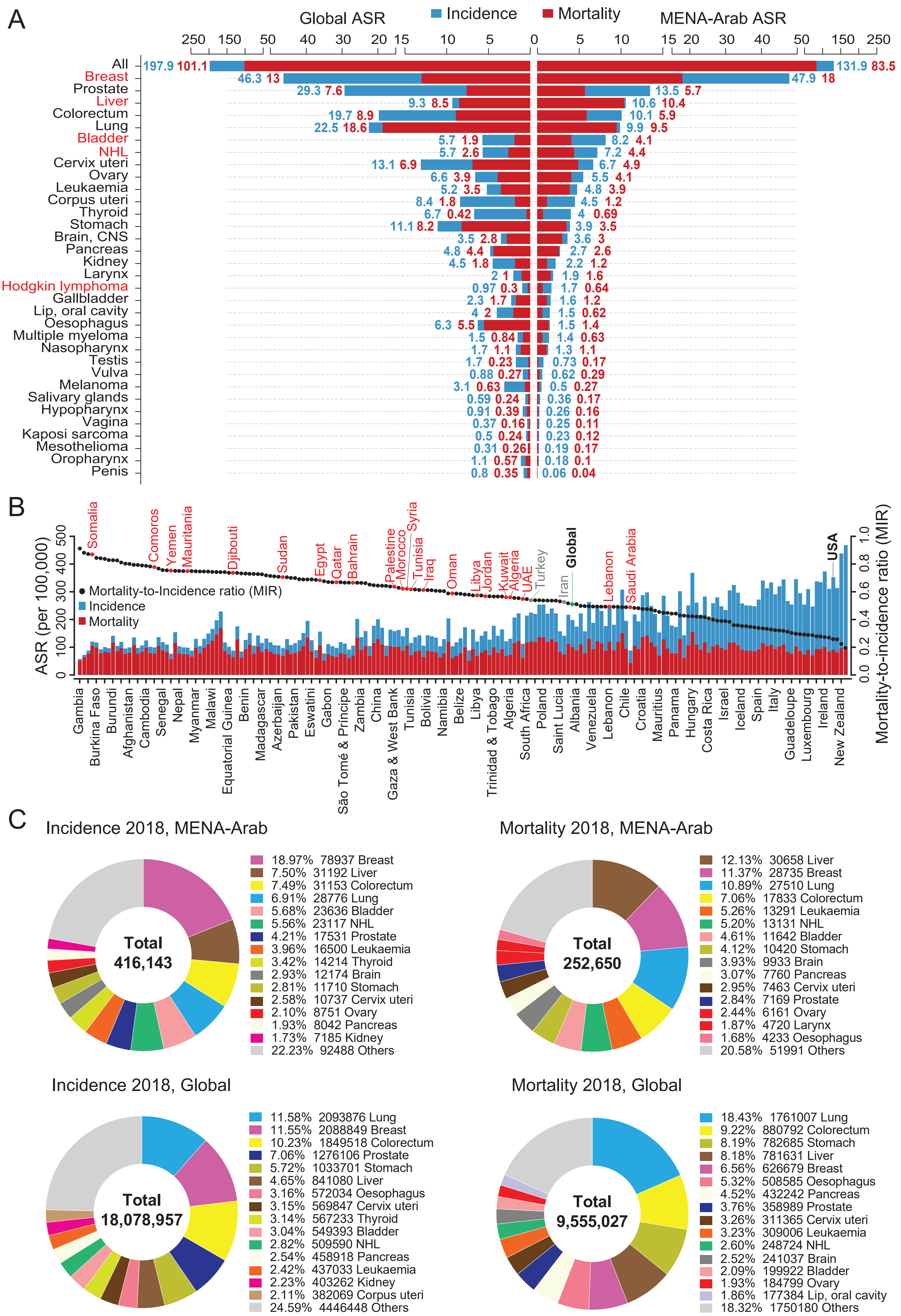


**Figure S1: Age-standardized rate (ASR) for cancer incidence and mortality, and mortality-to-incidence ratio (MIR) in both sexes in 2018.** (A) Comparison of ASR (per 100,000 people) in Arab countries in the MENA region (right) to the worldwide (global) ASR (left) for cancer incidence (blue) and cancer mortality (red) in 2018 for all ages. ASR is shown for all cancers and each cancer site. Cancer sites are ranked according to ASR incidence in the MENA-Arab region. The ASR for incidence and mortality are shown in blue and red, respectively. Cancer sites that are labelled in red had higher incidence and mortality in the MENA-Arab region than the global rates in 2018. (B) ASR for incidence (blue) and mortality (red) for all cancers in 2018 in both sexes for all ages in 186 countries. Countries were ranked according to the mortality-to-incidence ratio (MIR); mortality ASR divided by incidence ASR (black dots). The 22 Arab countries are labelled in red. (C) Top 15 cancer sites for incidence (B) and mortality (C) in both sexes, in 2018 for all ages, in the MENA region and worldwide. The total number of cases is shown in the middle of the donut graphs; the percentage contribution and the number of cases of each cancer site are shown in the legends.

#### Supplementary Figure 2

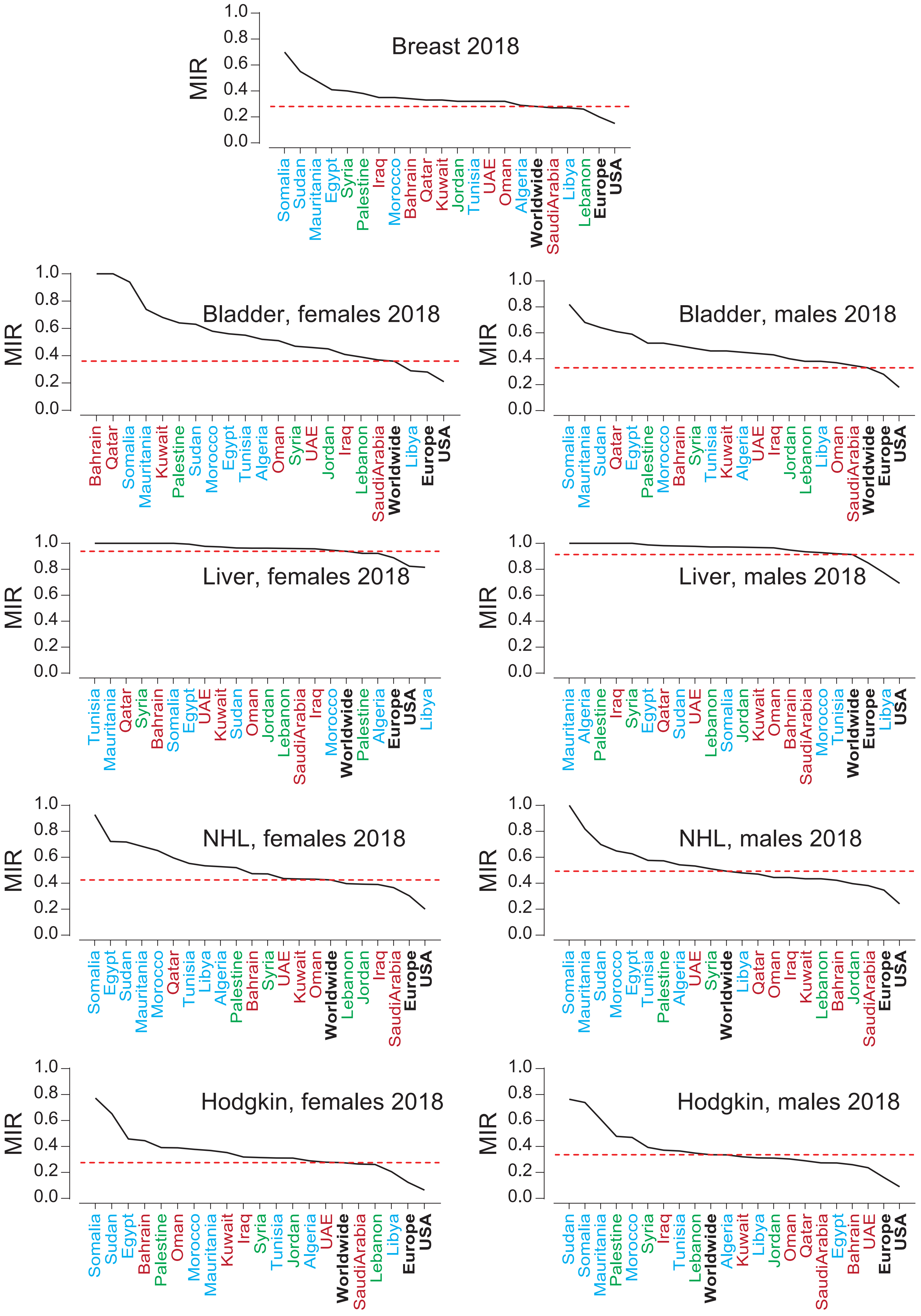


**Figure S2: Mortality-to-incidence ratio (MIR) for five cancers with higher incidence in the Arab region**. The MIR was calculated as the ratio of ASR for mortality to the ASR for incidence for all ages. The dotted lines mark the worldwide MIR. Arab countries are colored as those in the Arabian Gulf (maroon), the Levant (green) and North Africa (blue).

**Supplementary Figure 3**

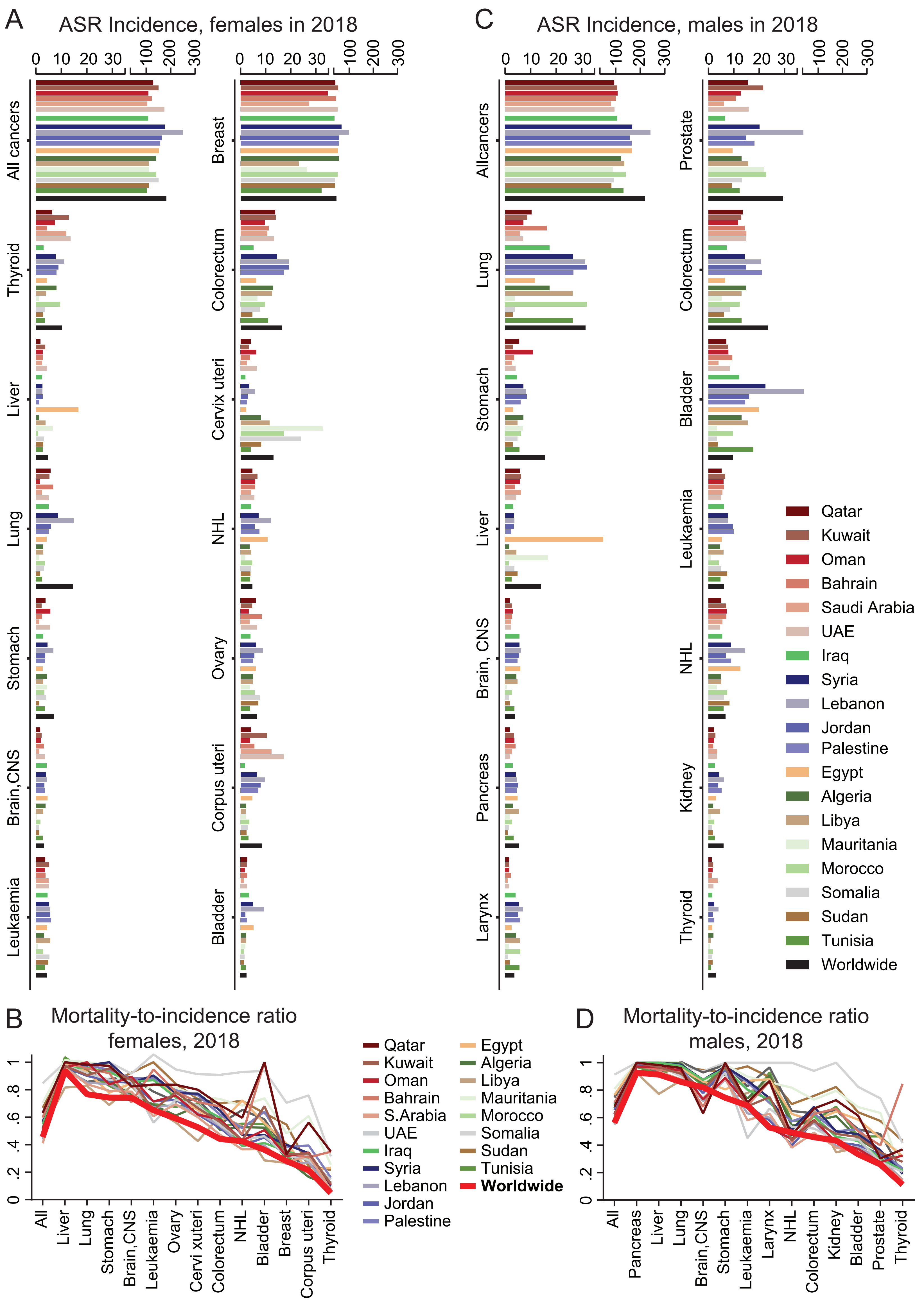


**Figure S3: The top cancer sites in the MENA-Arab region leading incidence or mortality in females or males in 2018 for all ages**. The MENA-Arab and worldwide ASR in 2018 for incidence for leading cancer sites for all ages in the MENA-Arab countries (coloured bars) and the worldwide rate for these sites (black bars) for females (A) and males (C). Mortality-to-incidence ratio (MIR) for the leading cancer sites in the MENA-Arab countries and the world for females (B) and males (D). Complete data mortality ASRs are available in Tables S1-S2.

**Supplementary Figure 4**

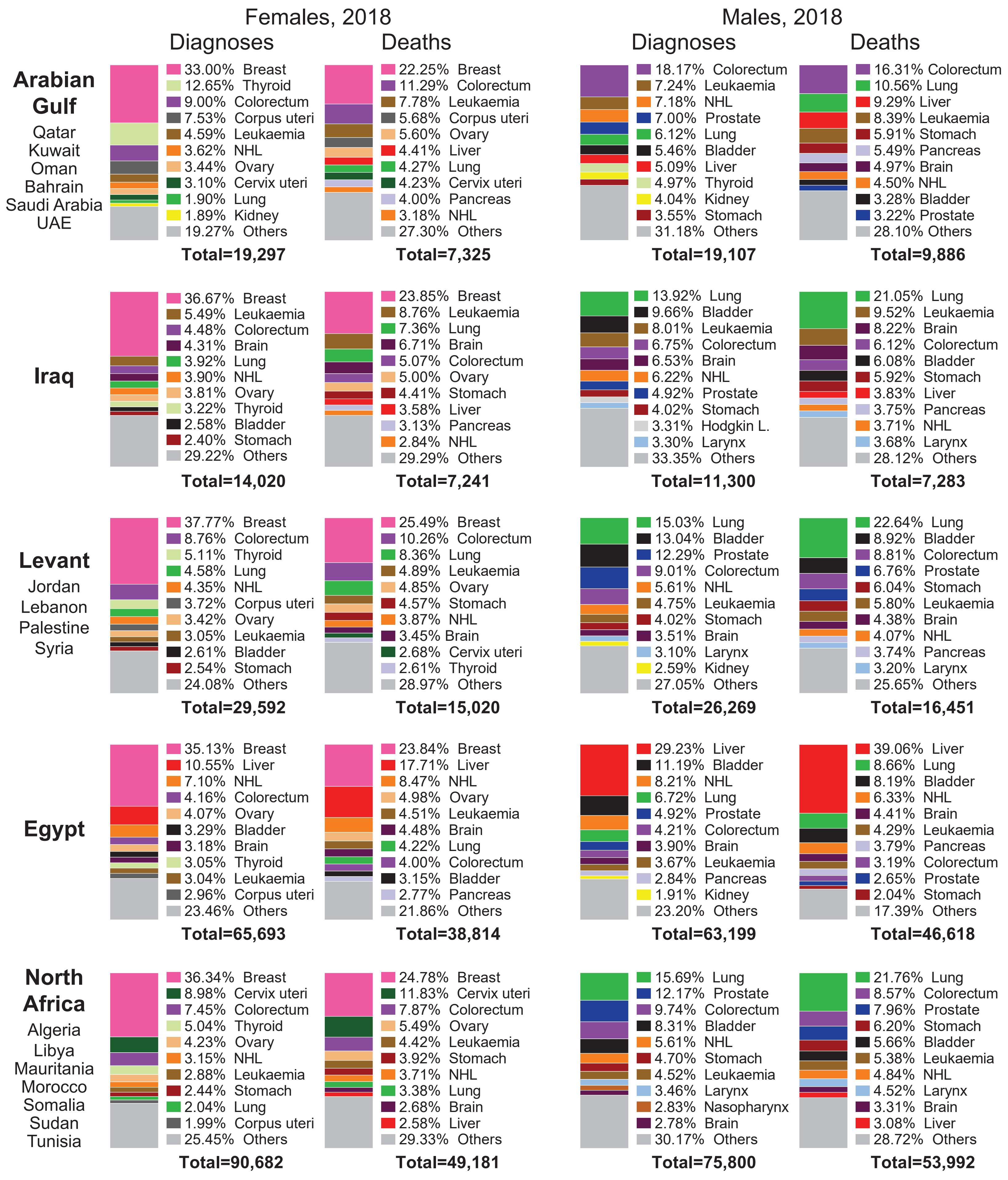


**Figure S4: Leading cancers for diagnosis and cancer-related deaths in subregions of the Arab countries**. The percentage contribution of cancer sites to all cancers in females or males are shown for the 10 leading sites for cancer diagnoses and mortality in 2018 at all ages in the specified Arab subregions. The total number of diagnoses or deaths are shown under each plot. Complete data for all cancer sites in all subregions, in each country, and data for the USA, Europe and the world are available in Tables S1-S2.

#### Supplementary Figure 5

**Figure S5: Age-specific incidence of cancers in Arab countries**. The ASIR for cancers at different age groups in Arab countries are summarized and compared to the ASIR at the same age groups for the World, US and Europe. Arab countries are labelled as those in the Arabian Gulf (maroon), the Levant (green) and North Africa (blue). The world ASIR is marked with bold font, USA and Europe in black font. Each cancer type is shown on a separate page in the following pages. For all age groups please refer to **Supplementary Data**.

### Bladder

**Females**

| 20-54 | | 55-69 | | 70-85+ | |
|---|---|---|---|---|---|
| Kuwait | 0.19 | S.Arabia | 4.8 | Mauritania | 9.8 |
| UAE | 0.19 | Morocco | 5.1 | Sudan | 11.4 |
| Bahrain | 0.39 | Jordan | 5.2 | S.Arabia | 12.0 |
| Qatar | 0.41 | Palestine | 5.3 | Somalia | 12.7 |
| Morocco | 0.42 | Oman | 5.7 | Morocco | 14.4 |
| Tunisia | 0.51 | Sudan | 5.9 | Libya | 17.2 |
| S.Arabia | 0.59 | Tunisia | 6.6 | Oman | 19.1 |
| Oman | 0.63 | Somalia | 7.3 | UAE | 20.1 |
| Libya | 0.65 | Mauritania | 7.5 | Jordan | 21.9 |
| Algeria | 0.66 | Algeria | 7.8 | Algeria | 23.6 |
| Palestine | 0.68 | Bahrain | 8.0 | **Worldwide** | 26.0 |
| Sudan | 0.72 | **Worldwide** | 8.7 | Tunisia | 26.5 |
| Somalia | 0.75 | Kuwait | 9.3 | Iraq | 26.6 |
| **Worldwide** | 0.85 | Libya | 10.0 | Qatar | 28.0 |
| Jordan | 0.86 | Qatar | 12.2 | Kuwait | 33.8 |
| Iraq | 1.10 | UAE | 14.6 | Bahrain | 38.1 |
| Egypt | 1.40 | Iraq | 16.3 | Palestine | 39.7 |
| Mauritania | 1.40 | Europe | 17.0 | Europe | 41.2 |
| USA | 1.60 | Syria | 19.2 | Syria | 45.6 |
| Europe | 1.80 | USA | 20.6 | Egypt | 49.1 |
| Syria | 2.00 | Egypt | 24.0 | USA | 54.8 |
| Lebanon | 4.10 | Lebanon | 37.0 | Lebanon | 86.5 |

**Males**

| 20-54 | | 55-69 | | 70-85+ | |
|---|---|---|---|---|---|
| UAE | 0.56 | Sudan | 11.7 | Somalia | 25.3 |
| Qatar | 1.10 | Mauritania | 12.2 | Mauritania | 30.8 |
| Kuwait | 1.30 | Somalia | 13.1 | Sudan | 31.1 |
| Bahrain | 1.40 | S.Arabia | 15.7 | S.Arabia | 33.6 |
| Mauritania | 1.90 | Qatar | 22.3 | Oman | 85.8 |
| Somalia | 2.00 | Kuwait | 24.0 | Morocco | 100.0 |
| S.Arabia | 2.10 | Oman | 26.6 | Qatar | 100.4 |
| **Worldwide** | 2.30 | UAE | 30.0 | Iraq | 107.7 |
| Sudan | 2.40 | Bahrain | 34.8 | Kuwait | 109.8 |
| Morocco | 2.70 | **Worldwide** | 35.8 | **Worldwide** | 114.1 |
| Oman | 3.30 | Morocco | 41.2 | UAE | 119.1 |
| Algeria | 3.50 | Palestine | 41.6 | Bahrain | 122.6 |
| Iraq | 3.60 | Libya | 55.1 | Algeria | 126.4 |
| Libya | 3.70 | Iraq | 56.1 | Jordan | 179.7 |
| Palestine | 4.20 | Jordan | 56.6 | Tunisia | 189.6 |
| USA | 4.60 | Algeria | 57.7 | Egypt | 191.7 |
| Europe | 5.00 | Tunisia | 67.1 | Libya | 194.6 |
| Jordan | 5.80 | USA | 76.9 | Palestine | 200.4 |
| Tunisia | 6.00 | Europe | 83.5 | Europe | 218.6 |
| Egypt | 6.40 | Egypt | 83.9 | Syria | 228.5 |
| Syria | 7.80 | Syria | 89.4 | USA | 236.7 |
| Lebanon | 15.90 | Lebanon | 153.6 | Lebanon | 397.9 |

### Brain, CNS

#### Females

| 0-14 | | 15-29 | | 30-54 | | 55-69 | | 70-85+ | |
|---|---|---|---|---|---|---|---|---|---|
| Mauritania | 0.23 | Qatar | | Mauritania | 0.92 | Mauritania | 0.83 | Mauritania | |
| Somalia | 0.37 | Mauritania | | UAE | 0.95 | Sudan | 3.30 | Qatar | |
| Sudan | 0.53 | Oman | 0.18 | Somalia | 1.20 | S.Arabia | 3.40 | Sudan | 3.1 |
| S.Arabia | 0.93 | Kuwait | 0.21 | Bahrain | 1.40 | Somalia | 4.50 | Somalia | 3.2 |
| Morocco | 0.97 | UAE | 0.32 | S.Arabia | 1.40 | Oman | 4.50 | Morocco | 3.5 |
| Oman | 0.97 | Somalia | 0.35 | Sudan | 1.80 | Morocco | 5.60 | S.Arabia | 4.1 |
| **Worldwide** | 1.10 | Sudan | 0.36 | Kuwait | 2.20 | Tunisia | 6.50 | Oman | 5.5 |
| Iraq | 1.10 | Morocco | 0.44 | Morocco | 2.20 | Qatar | 6.70 | Libya | 8.0 |
| Kuwait | 1.10 | Bahrain | 0.51 | Qatar | 2.20 | Kuwait | 7.00 | Tunisia | 8.1 |
| Qatar | 1.10 | S.Arabia | 0.63 | Libya | 2.70 | Palestine | 7.60 | Algeria | 8.7 |
| Tunisia | 1.20 | **Worldwide** | 1.00 | Oman | 2.70 | **Worldwide** | 8.90 | Bahrain | 11.4 |
| Jordan | 1.40 | Jordan | 1.10 | Tunisia | 3.20 | Libya | 9.00 | Jordan | 11.5 |
| Syria | 1.40 | Libya | 1.20 | **Worldwide** | 3.30 | Algeria | 9.60 | Kuwait | 12.2 |
| Libya | 1.50 | Palestine | 1.20 | Palestine | 3.50 | Jordan | 10.30 | **Worldwide** | 13.3 |
| Algeria | 1.80 | Iraq | 1.30 | Jordan | 3.60 | Lebanon | 10.40 | Syria | 14.1 |
| Lebanon | 1.80 | Tunisia | 1.30 | USA | 4.20 | Syria | 10.50 | Iraq | 14.3 |
| Egypt | 1.90 | Syria | 1.50 | Egypt | 4.70 | USA | 10.90 | Lebanon | 16.6 |
| Palestine | 1.90 | Algeria | 1.80 | Algeria | 4.90 | Iraq | 11.90 | USA | 16.7 |
| UAE | 1.90 | Egypt | 1.80 | Europe | 4.90 | Bahrain | 12.00 | Palestine | 17.7 |
| Europe | 2.20 | Europe | 1.80 | Syria | 4.90 | Egypt | 12.90 | Europe | 17.8 |
| Bahrain | 2.70 | Lebanon | 2.00 | Lebanon | 5.10 | Europe | 12.90 | Egypt | 19.9 |
| USA | 3.40 | USA | 2.30 | Iraq | 5.70 | UAE | 13.70 | UAE | 28.9 |

#### Males

| 0-14 | | 15-29 | | 30-54 | | 55-69 | | 70-85+ | |
|---|---|---|---|---|---|---|---|---|---|
| Mauritania | 0.33 | Mauritania | 0.15 | Mauritania | 0.76 | Mauritania | 1.6 | Mauritania | 2.5 |
| Sudan | 0.36 | UAE | 0.51 | UAE | 1.20 | Sudan | 3.6 | Morocco | 5.9 |
| Somalia | 0.48 | Sudan | 0.52 | Sudan | 1.50 | Qatar | 4.9 | Oman | 6.1 |
| Qatar | 1.00 | Somalia | 0.59 | Kuwait | 1.60 | Somalia | 4.9 | Somalia | 6.9 |
| Bahrain | 1.30 | Kuwait | 0.60 | Qatar | 1.60 | S.Arabia | 5.8 | Tunisia | 7.0 |
| **Worldwide** | 1.30 | Qatar | 0.79 | Somalia | 1.80 | Morocco | 6.8 | UAE | 8.1 |
| Kuwait | 1.30 | Oman | 0.80 | Bahrain | 1.90 | Oman | 8.0 | S.Arabia | 8.9 |
| Libya | 1.40 | S.Arabia | 0.86 | S.Arabia | 2.10 | Kuwait | 8.5 | Bahrain | 9.6 |
| S.Arabia | 1.40 | Morocco | 0.97 | Morocco | 3.40 | UAE | 9.7 | Qatar | 10.7 |
| Iraq | 1.50 | Tunisia | 0.98 | Oman | 3.80 | Tunisia | 9.9 | Libya | 11.0 |
| UAE | 1.50 | Bahrain | 1.10 | **Worldwide** | 4.10 | Bahrain | 10.8 | Algeria | 12.4 |
| Oman | 1.90 | Libya | 1.10 | Tunisia | 4.60 | **Worldwide** | 11.5 | Lebanon | 12.8 |
| Egypt | 2.00 | Palestine | 1.20 | Algeria | 5.30 | Algeria | 13.0 | Kuwait | 16.2 |
| Syria | 2.00 | Algeria | 1.30 | Palestine | 5.50 | Palestine | 13.0 | Syria | 16.2 |
| Algeria | 2.10 | **Worldwide** | 1.30 | Egypt | 5.70 | Egypt | 15.6 | Jordan | 16.8 |
| Morocco | 2.10 | Iraq | 1.40 | USA | 6.00 | Lebanon | 15.6 | **Worldwide** | 17.8 |
| Tunisia | 2.20 | Jordan | 1.60 | Libya | 6.30 | Syria | 15.7 | Sudan | 19.7 |
| Jordan | 2.40 | Syria | 1.80 | Jordan | 6.80 | Jordan | 15.9 | Iraq | 21.0 |
| Palestine | 2.40 | Egypt | 2.10 | Iraq | 7.20 | USA | 16.0 | Palestine | 22.2 |
| Europe | 2.50 | Europe | 2.50 | Europe | 7.30 | Iraq | 16.7 | USA | 24.2 |
| Lebanon | 2.50 | Lebanon | 2.60 | Syria | 7.50 | Libya | 17.3 | Europe | 25.7 |
| USA | 3.80 | USA | 2.80 | Lebanon | 8.60 | Europe | 19.1 | Egypt | 33.8 |

#### Breast

| 20-34 | | 35-49 | | 50-69 | | 70-85+ | |
|---|---|---|---|---|---|---|---|
| Qatar | 5.3 | Mauritania | 46.3 | Libya | 64.5 | Libya | 48.7 |
| Kuwait | 5.4 | Libya | 48.4 | Mauritania | 78.4 | S.Arabia | 69.4 |
| Bahrain | 7.0 | S.Arabia | 52.4 | S.Arabia | 78.9 | Oman | 73.9 |
| UAE | 7.7 | Qatar | 55.6 | Tunisia | 89.6 | Mauritania | 84.1 |
| Mauritania | 9.8 | Tunisia | 60.1 | Sudan | 98.4 | Iraq | 90.4 |
| Libya | 10.2 | Kuwait | 63.8 | Oman | 102.3 | Tunisia | 94.2 |
| S.Arabia | 11.2 | Bahrain | 69.1 | Somalia | 110.9 | Algeria | 108.4 |
| Iraq | 12.5 | Oman | 69.2 | Iraq | 116.5 | Morocco | 116.2 |
| **Worldwide** | 12.6 | **Worldwide** | 72.1 | **Worldwide** | 145.1 | Somalia | 118.8 |
| Oman | 13.2 | Sudan | 73.4 | Algeria | 149.8 | Qatar | 140.5 |
| Jordan | 14.6 | Iraq | 74.2 | Bahrain | 150.5 | Sudan | 148.1 |
| Tunisia | 14.6 | UAE | 74.6 | Morocco | 155.5 | Bahrain | 150.4 |
| Palestine | 14.8 | Somalia | 75.8 | Egypt | 157.2 | **Worldwide** | 182.7 |
| USA | 15.5 | Egypt | 87.7 | Qatar | 158.3 | Jordan | 183.7 |
| Egypt | 15.6 | Palestine | 93.9 | Palestine | 163.6 | Egypt | 200.6 |
| Europe | 16.6 | Morocco | 98.6 | UAE | 177.6 | Syria | 219.6 |
| Morocco | 16.6 | Jordan | 101.9 | Jordan | 178.4 | UAE | 234.2 |
| Sudan | 19.3 | Europe | 114.3 | Syria | 191.3 | Kuwait | 241.4 |
| Somalia | 23.2 | USA | 115.5 | Kuwait | 193.9 | Palestine | 255.6 |
| Algeria | 26.6 | Algeria | 118.9 | Europe | 239.9 | Europe | 293.9 |
| Syria | 27.1 | Syria | 121.8 | Lebanon | 274.2 | Lebanon | 366.5 |
| Lebanon | 28.8 | Lebanon | 181.2 | USA | 282.3 | USA | 387.0 |

### Cervix uteri

| 15-34 | | 35-54 | | 55-69 | | 70-85+ | |
|---|---|---|---|---|---|---|---|
| Bahrain | | Egypt | 3.7 | Palestine | 5.5 | Bahrain | |
| Libya | 0.14 | Iraq | 4.1 | Iraq | 5.9 | Iraq | 4.4 |
| Iraq | 0.29 | S.Arabia | 4.9 | Egypt | 7.5 | S.Arabia | 9.5 |
| Jordan | 0.29 | Palestine | 5.4 | S.Arabia | 8.3 | USA | 11.2 |
| Egypt | 0.30 | Jordan | 5.6 | Jordan | 8.6 | Kuwait | 12.2 |
| Palestine | 0.35 | Kuwait | 6.3 | Syria | 10.1 | Egypt | 12.5 |
| S.Arabia | 0.38 | Tunisia | 6.4 | Kuwait | 10.9 | Jordan | 13.1 |
| Qatar | 0.43 | Qatar | 7.0 | USA | 12.7 | Syria | 14.0 |
| Algeria | 0.44 | Bahrain | 7.2 | Qatar | 14.6 | Palestine | 14.2 |
| UAE | 0.46 | Syria | 7.2 | Tunisia | 15.2 | Qatar | 15.2 |
| Tunisia | 0.58 | UAE | 9.0 | Lebanon | 16.4 | Tunisia | 16.1 |
| Syria | 0.64 | Lebanon | 11.9 | Bahrain | 19.4 | Europe | 17.2 |
| Kuwait | 0.65 | Oman | 13.1 | Oman | 19.7 | Oman | 19.1 |
| Sudan | 0.84 | Algeria | 13.5 | Europe | 23.0 | Lebanon | 22.5 |
| Lebanon | 0.97 | USA | 13.9 | UAE | 24.5 | Sudan | 27.2 |
| Oman | 1.00 | Sudan | 14.5 | Sudan | 31.5 | **Worldwide** | 29.8 |
| Morocco | 2.10 | Libya | 19.2 | Algeria | 32.1 | Algeria | 32.2 |
| **Worldwide** | 4.50 | Europe | 24.8 | **Worldwide** | 35.6 | UAE | 36.9 |
| USA | 4.80 | **Worldwide** | 28.5 | Libya | 47.2 | Libya | 46.4 |
| Mauritania | 4.90 | Morocco | 30.4 | Morocco | 62.6 | Somalia | 64.9 |
| Somalia | 4.90 | Somalia | 48.0 | Somalia | 80.1 | Morocco | 66.0 |
| Europe | 7.30 | Mauritania | 53.8 | Mauritania | 116.5 | Mauritania | 153.8 |

#### Colorectum

##### Females

| 15-34 | | 35-54 | | 55-69 | | 70-85+ | |
|---|---|---|---|---|---|---|---|
| Qatar | 0.21 | Mauritania | 6.0 | Sudan | 15.2 | Sudan | 20.5 |
| Kuwait | 0.63 | Iraq | 6.5 | Iraq | 19.9 | Somalia | 25.3 |
| Mauritania | 0.64 | Sudan | 8.2 | Egypt | 21.0 | Iraq | 33.0 |
| Bahrain | 0.73 | Egypt | 8.4 | Mauritania | 24.2 | Egypt | 42.0 |
| Oman | 0.77 | Qatar | 9.8 | Somalia | 30.2 | Morocco | 48.5 |
| Iraq | 0.80 | Kuwait | 10.3 | Oman | 34.7 | Libya | 58.6 |
| UAE | 0.85 | Tunisia | 11.8 | S.Arabia | 39.0 | S.Arabia | 60.6 |
| Sudan | 0.99 | Bahrain | 12.1 | Morocco | 41.0 | Mauritania | 62.0 |
| Morocco | 1.00 | Somalia | 12.6 | Tunisia | 42.1 | Oman | 63.6 |
| Palestine | 1.00 | Oman | 12.9 | Libya | 42.7 | UAE | 73.0 |
| Syria | 1.00 | Morocco | 13.1 | Bahrain | 47.5 | Bahrain | 73.1 |
| **Worldwide** | 1.10 | **Worldwide** | 14.0 | Algeria | 50.7 | Algeria | 76.0 |
| Jordan | 1.10 | S.Arabia | 15.4 | UAE | 52.8 | Tunisia | 78.0 |
| Lebanon | 1.10 | Algeria | 17.1 | Syria | 53.0 | Syria | 107.2 |
| S.Arabia | 1.10 | Syria | 17.9 | Palestine | 60.5 | Kuwait | 114.3 |
| Egypt | 1.20 | UAE | 19.5 | **Worldwide** | 60.7 | Qatar | 115.4 |
| Somalia | 1.30 | Libya | 20.8 | Kuwait | 62.0 | Jordan | 125.9 |
| Europe | 1.40 | Palestine | 20.8 | Qatar | 62.6 | Palestine | 137.8 |
| Algeria | 1.50 | Europe | 21.1 | Lebanon | 63.7 | **Worldwide** | 150.4 |
| Tunisia | 1.60 | Lebanon | 23.3 | Jordan | 74.4 | Lebanon | 162.0 |
| USA | 2.10 | Jordan | 24.6 | USA | 80.9 | USA | 176.0 |
| Libya | 2.20 | USA | 26.3 | Europe | 96.6 | Europe | 205.4 |

##### Males

| 15-34 | | 35-54 | | 55-69 | | 70-85+ | |
|---|---|---|---|---|---|---|---|
| Qatar | 0.37 | Kuwait | 5.6 | Mauritania | 19.3 | Mauritania | 32.4 |
| UAE | 0.42 | Qatar | 5.7 | Sudan | 20.1 | Egypt | 38.9 |
| Bahrain | 0.46 | Bahrain | 6.0 | Egypt | 22.0 | Sudan | 43.7 |
| Libya | 0.54 | Mauritania | 6.3 | Iraq | 27.6 | Iraq | 45.7 |
| Jordan | 0.67 | UAE | 6.4 | Somalia | 35.0 | Somalia | 51.1 |
| Kuwait | 0.72 | Sudan | 7.9 | Oman | 41.7 | Oman | 63.4 |
| Palestine | 0.98 | Iraq | 8.5 | Qatar | 44.7 | Morocco | 77.7 |
| **Worldwide** | 1.00 | Egypt | 9.4 | Syria | 49.5 | Libya | 87.5 |
| Somalia | 1.00 | Somalia | 9.6 | Kuwait | 52.9 | Tunisia | 96.0 |
| Iraq | 1.10 | Morocco | 11.5 | Tunisia | 53.6 | S.Arabia | 102.1 |
| Tunisia | 1.10 | Tunisia | 12.7 | Morocco | 54.5 | Algeria | 112.9 |
| Algeria | 1.20 | Libya | 13.1 | Jordan | 55.3 | Syria | 125.0 |
| Mauritania | 1.20 | Jordan | 14.0 | Libya | 57.6 | Jordan | 133.1 |
| S.Arabia | 1.20 | Algeria | 14.2 | Bahrain | 59.3 | UAE | 136.9 |
| Sudan | 1.20 | Syria | 14.9 | Algeria | 60.7 | Kuwait | 139.9 |
| Syria | 1.20 | S.Arabia | 16.2 | S.Arabia | 61.3 | Bahrain | 154.4 |
| Europe | 1.30 | **Worldwide** | 16.6 | Lebanon | 66.7 | Qatar | 178.2 |
| Morocco | 1.30 | Oman | 17.6 | UAE | 69.9 | Lebanon | 194.8 |
| Egypt | 1.60 | Palestine | 18.8 | Palestine | 77.3 | Palestine | 200.2 |
| Oman | 1.60 | Lebanon | 22.6 | **Worldwide** | 94.9 | **Worldwide** | 224.2 |
| Lebanon | 1.70 | Europe | 23.0 | USA | 110.9 | USA | 226.0 |
| USA | 1.70 | USA | 30.4 | Europe | 161.6 | Europe | 349.5 |

### Corpus uteri

| 25-39 | | 40-54 | | 55-69 | | 70-85+ | |
|---|---|---|---|---|---|---|---|
| Libya | | Libya | 1.4 | Libya | 9.1 | Iraq | 5.3 |
| UAE | | Mauritania | 2.9 | Iraq | 9.1 | Bahrain | 6.5 |
| Morocco | 0.10 | Sudan | 2.9 | Mauritania | 11.3 | Algeria | 7.9 |
| Mauritania | 0.22 | Iraq | 3.0 | Algeria | 11.3 | Sudan | 11.0 |
| Egypt | 0.38 | Somalia | 3.2 | Sudan | 12.0 | Tunisia | 13.6 |
| Qatar | 0.39 | Oman | 3.2 | Somalia | 13.7 | Mauritania | 14.0 |
| Iraq | 0.43 | Algeria | 3.8 | Tunisia | 15.2 | Qatar | 15.2 |
| Tunisia | 0.43 | Morocco | 4.3 | Morocco | 19.7 | Morocco | 15.4 |
| Algeria | 0.44 | Tunisia | 5.0 | Oman | 21.2 | Oman | 17.7 |
| Sudan | 0.55 | Egypt | 5.7 | Qatar | 21.3 | Somalia | 18.3 |
| Bahrain | 0.56 | Qatar | 6.4 | Egypt | 23.2 | Libya | 18.8 |
| Jordan | 0.62 | S.Arabia | 7.6 | Bahrain | 27.2 | Egypt | 28.1 |
| Somalia | 0.67 | Palestine | 8.6 | Syria | 28.9 | Syria | 36.0 |
| Kuwait | 0.72 | Kuwait | 8.8 | Palestine | 31.0 | **Worldwide** | 39.6 |
| Palestine | 0.79 | Jordan | 9.8 | Jordan | 36.0 | Lebanon | 50.7 |
| Oman | 0.96 | Syria | 9.8 | **Worldwide** | 36.7 | Palestine | 51.9 |
| Syria | 0.98 | UAE | 10.3 | Lebanon | 41.3 | Jordan | 52.9 |
| S.Arabia | 1.10 | Bahrain | 12.6 | Kuwait | 53.7 | UAE | 67.6 |
| Europe | 1.70 | **Worldwide** | 14.2 | S.Arabia | 54.7 | Kuwait | 70.2 |
| **Worldwide** | 1.70 | Lebanon | 15.9 | Europe | 75.9 | Europe | 78.8 |
| Lebanon | 1.80 | Europe | 23.5 | USA | 96.6 | USA | 94.3 |
| USA | 2.80 | USA | 30.0 | UAE | 116.7 | S.Arabia | 119.0 |

### Gallbladder

#### Females

| 30-54 | | 55-69 | | 70-85+ | |
|---|---|---|---|---|---|
| Mauritania | 0.22 | Egypt | 3.4 | Egypt | 5.5 |
| Qatar | 0.53 | Oman | 3.4 | Mauritania | 7.7 |
| UAE | 0.67 | Mauritania | 3.9 | Somalia | 7.7 |
| Europe | 0.75 | Sudan | 4.8 | Sudan | 8.6 |
| Kuwait | 0.75 | S.Arabia | 5.2 | Oman | 10.4 |
| Iraq | 0.86 | Jordan | 5.8 | Morocco | 10.5 |
| Egypt | 0.91 | Palestine | 6.1 | Bahrain | 11.4 |
| S.Arabia | 0.91 | USA | 6.5 | S.Arabia | 11.6 |
| Sudan | 0.91 | Europe | 6.6 | Iraq | 16.8 |
| USA | 0.93 | Lebanon | 6.9 | USA | 16.9 |
| Bahrain | 1.20 | Somalia | 7.2 | Europe | 19.0 |
| Morocco | 1.30 | Syria | 7.4 | Palestine | 19.8 |
| Oman | 1.30 | Bahrain | 8.0 | Kuwait | 19.8 |
| Syria | 1.40 | Morocco | 8.0 | Lebanon | 20.8 |
| Lebanon | 1.50 | Kuwait | 8.5 | **Worldwide** | 23.3 |
| Palestine | 1.50 | **Worldwide** | 8.6 | Syria | 24.8 |
| **Worldwide** | 1.60 | Tunisia | 8.6 | Tunisia | 25.7 |
| Tunisia | 1.60 | Iraq | 9.0 | Qatar | 28.0 |
| Somalia | 1.80 | Libya | 10.5 | Algeria | 29.3 |
| Jordan | 1.90 | Qatar | 12.2 | Jordan | 35.3 |
| Libya | 2.30 | UAE | 16.2 | Libya | 44.1 |
| Algeria | 3.40 | Algeria | 20.8 | UAE | 46.3 |

#### Males

| 30-54 | | 55-69 | | 70-85+ | |
|---|---|---|---|---|---|
| Mauritania | | Mauritania | | Mauritania | |
| Sudan | 0.20 | Sudan | 1.7 | Sudan | 3.7 |
| Qatar | 0.27 | Somalia | 2.5 | Morocco | 6.6 |
| UAE | 0.36 | Egypt | 3.4 | Somalia | 6.7 |
| Bahrain | 0.37 | Morocco | 4.2 | Iraq | 6.9 |
| Somalia | 0.50 | Iraq | 4.4 | Egypt | 9.6 |
| Oman | 0.52 | S.Arabia | 4.6 | Bahrain | 9.6 |
| Kuwait | 0.58 | Qatar | 4.9 | Lebanon | 12.1 |
| Palestine | 0.58 | Bahrain | 5.9 | S.Arabia | 13.0 |
| Libya | 0.63 | Kuwait | 6.1 | Tunisia | 15.3 |
| Morocco | 0.68 | Syria | 6.3 | Libya | 15.7 |
| S.Arabia | 0.68 | UAE | 6.6 | Syria | 16.0 |
| Iraq | 0.70 | Libya | 6.7 | Algeria | 17.2 |
| Tunisia | 0.79 | Palestine | 6.8 | USA | 18.2 |
| Egypt | 0.80 | Lebanon | 7.0 | Kuwait | 21.3 |
| Europe | 0.80 | USA | 7.0 | Palestine | 22.1 |
| Syria | 0.93 | Jordan | 7.2 | Europe | 22.4 |
| **Worldwide** | 1.00 | Tunisia | 7.4 | **Worldwide** | 24.8 |
| Lebanon | 1.00 | Europe | 7.6 | Jordan | 27.9 |
| USA | 1.00 | **Worldwide** | 8.2 | Oman | 28.6 |
| Jordan | 1.20 | Algeria | 8.9 | Qatar | 33.5 |
| Algeria | 1.40 | Oman | 14.2 | UAE | 54.7 |

#### Hodgkin Lymphoma

##### Females

| 0-14 | | 15-34 | | 35-54 | | 55-69 | | 70-85+ | |
|---|---|---|---|---|---|---|---|---|---|
| Bahrain | | Mauritania | 0.13 | Bahrain | | Mauritania | | Mauritania | |
| Mauritania | 0.11 | Sudan | 0.39 | Qatar | | Qatar | | Qatar | |
| Egypt | 0.24 | Somalia | 0.48 | Mauritania | 0.52 | Sudan | 0.70 | Somalia | |
| **Worldwide** | 0.26 | Bahrain | 0.86 | **Worldwide** | 0.85 | Somalia | 0.83 | Libya | |
| Iraq | 0.34 | Egypt | 0.90 | Egypt | 0.97 | **Worldwide** | 1.20 | Bahrain | |
| UAE | 0.35 | **Worldwide** | 0.98 | Somalia | 0.99 | Libya | 1.60 | UAE | |
| Oman | 0.39 | Oman | 1.80 | Sudan | 1.30 | Morocco | 1.60 | Sudan | 1.3 |
| Libya | 0.41 | Iraq | 1.90 | Kuwait | 1.30 | Europe | 1.90 | **Worldwide** | 2.2 |
| Sudan | 0.48 | Qatar | 1.90 | S.Arabia | 1.40 | Algeria | 2.00 | Europe | 2.4 |
| Qatar | 0.53 | UAE | 2.00 | UAE | 1.50 | Kuwait | 2.10 | Oman | 3.2 |
| Syria | 0.55 | Kuwait | 2.10 | Jordan | 1.50 | Tunisia | 2.10 | USA | 3.3 |
| Algeria | 0.59 | S.Arabia | 2.40 | Tunisia | 1.50 | Egypt | 2.30 | Iraq | 3.4 |
| Morocco | 0.61 | Morocco | 2.50 | Morocco | 1.80 | Jordan | 2.30 | Algeria | 3.5 |
| Palestine | 0.61 | Jordan | 2.70 | Palestine | 2.10 | USA | 2.40 | Tunisia | 3.5 |
| USA | 0.62 | Libya | 2.70 | Iraq | 2.20 | Iraq | 2.70 | Kuwait | 3.7 |
| Europe | 0.63 | Syria | 2.70 | Algeria | 2.20 | S.Arabia | 2.70 | S.Arabia | 3.8 |
| Somalia | 0.63 | Tunisia | 2.70 | Europe | 2.20 | Syria | 2.80 | Jordan | 4.7 |
| Kuwait | 0.68 | Algeria | 2.80 | USA | 2.40 | Palestine | 3.00 | Syria | 4.8 |
| Jordan | 0.72 | Palestine | 2.90 | Libya | 2.60 | Lebanon | 3.10 | Morocco | 6.5 |
| Lebanon | 0.75 | USA | 3.50 | Syria | 2.60 | Oman | 4.50 | Lebanon | 7.4 |
| Tunisia | 0.79 | Europe | 3.60 | Oman | 2.80 | Bahrain | 6.50 | Palestine | 8.4 |
| S.Arabia | 0.81 | Lebanon | 4.20 | Lebanon | 4.00 | UAE | 8.00 | Egypt | 8.8 |

##### Males

| 0-14 | | 15-34 | | 35-54 | | 55-69 | | 70-85+ | |
|---|---|---|---|---|---|---|---|---|---|
| Mauritania | 0.32 | Mauritania | 0.63 | UAE | 0.27 | Mauritania | 0.63 | Qatar | |
| **Worldwide** | 0.52 | Sudan | 0.90 | Mauritania | 0.41 | Palestine | 0.63 | Mauritania | |
| Qatar | 0.53 | Somalia | 0.96 | Bahrain | 0.68 | Libya | 0.90 | Bahrain | |
| Sudan | 0.55 | Bahrain | 1.10 | Somalia | 1.10 | Bahrain | 1.10 | Somalia | |
| Bahrain | 0.61 | **Worldwide** | 1.10 | **Worldwide** | 1.30 | Somalia | 1.10 | UAE | |
| Somalia | 0.65 | Libya | 1.50 | Sudan | 1.40 | **Worldwide** | 2.00 | Jordan | 1.7 |
| Libya | 0.70 | UAE | 1.50 | Qatar | 1.50 | Sudan | 2.30 | Sudan | 2.3 |
| UAE | 0.73 | Morocco | 1.70 | Kuwait | 1.60 | Jordan | 2.40 | **Worldwide** | 2.9 |
| USA | 0.76 | Egypt | 1.80 | Libya | 2.00 | S.Arabia | 2.40 | Algeria | 3.1 |
| Oman | 0.79 | Oman | 1.80 | Algeria | 2.10 | Kuwait | 2.70 | Kuwait | 3.5 |
| Kuwait | 0.85 | Jordan | 2.10 | Egypt | 2.20 | Egypt | 2.80 | Europe | 3.5 |
| Palestine | 0.87 | Iraq | 2.20 | S.Arabia | 2.30 | Algeria | 3.00 | Tunisia | 3.6 |
| Jordan | 0.88 | S.Arabia | 2.20 | Morocco | 2.40 | Europe | 3.10 | S.Arabia | 4.0 |
| Europe | 0.91 | Algeria | 2.30 | Iraq | 2.60 | Iraq | 3.10 | Oman | 4.1 |
| Morocco | 0.91 | Tunisia | 2.30 | Tunisia | 3.00 | UAE | 3.20 | USA | 4.8 |
| S.Arabia | 0.92 | Palestine | 2.50 | Europe | 3.00 | Qatar | 3.50 | Iraq | 5.1 |
| Tunisia | 0.96 | Kuwait | 2.90 | Palestine | 3.10 | Morocco | 3.60 | Morocco | 6.2 |
| Iraq | 1.00 | Syria | 2.90 | Jordan | 3.20 | USA | 3.70 | Egypt | 8.1 |
| Syria | 1.00 | Qatar | 3.20 | USA | 3.50 | Syria | 4.00 | Syria | 8.2 |
| Algeria | 1.10 | USA | 3.60 | Syria | 3.70 | Tunisia | 4.10 | Libya | 10.1 |
| Lebanon | 1.50 | Europe | 3.70 | Oman | 4.30 | Oman | 5.60 | Palestine | 11.5 |
| Egypt | 1.60 | Lebanon | 4.40 | Lebanon | 5.40 | Lebanon | 5.70 | Lebanon | 15.8 |

### Kidney

**Females**

| 20-39 | | 40-54 | | 55-69 | | 70-85+ | |
|---|---|---|---|---|---|---|---|
| Bahrain | | Kuwait | 1.2 | Mauritania | 1.4 | Mauritania | |
| Qatar | | Morocco | 1.5 | Oman | 3.3 | Bahrain | |
| Morocco | 0.15 | Algeria | 1.5 | Sudan | 3.9 | Sudan | 1.7 |
| Algeria | 0.26 | Mauritania | 1.5 | Morocco | 4.3 | Somalia | 2.4 |
| Tunisia | 0.32 | Bahrain | 1.8 | Algeria | 4.5 | Algeria | 4.0 |
| UAE | 0.32 | Egypt | 2.1 | Somalia | 4.6 | Oman | 5.5 |
| Libya | 0.38 | Sudan | 2.4 | Egypt | 5.3 | Morocco | 6.5 |
| Iraq | 0.39 | Libya | 2.5 | Bahrain | 5.6 | Iraq | 6.8 |
| Somalia | 0.39 | Iraq | 2.6 | Tunisia | 5.7 | Egypt | 7.1 |
| Sudan | 0.43 | Tunisia | 2.7 | Iraq | 5.8 | S.Arabia | 9.2 |
| Kuwait | 0.45 | UAE | 2.9 | Libya | 6.5 | Tunisia | 10.1 |
| Jordan | 0.46 | Somalia | 2.9 | Kuwait | 6.8 | Libya | 10.6 |
| Mauritania | 0.46 | Syria | 3.2 | Syria | 7.0 | Syria | 11.0 |
| Egypt | 0.51 | Lebanon | 3.2 | UAE | 7.5 | Kuwait | 11.8 |
| **Worldwide** | 0.53 | Jordan | 3.3 | Lebanon | 7.6 | Lebanon | 12.5 |
| S.Arabia | 0.54 | Palestine | 3.3 | Palestine | 7.6 | Qatar | 12.8 |
| Syria | 0.54 | **Worldwide** | 3.6 | S.Arabia | 7.9 | Jordan | 13.1 |
| Palestine | 0.55 | Qatar | 3.7 | Jordan | 8.0 | Palestine | 22.0 |
| Lebanon | 0.79 | S.Arabia | 3.8 | **Worldwide** | 11.7 | **Worldwide** | 22.0 |
| Oman | 0.79 | Oman | 3.9 | Qatar | 12.2 | UAE | 37.1 |
| Europe | 0.96 | Europe | 8.4 | Europe | 26.2 | Europe | 38.6 |
| USA | 1.70 | USA | 11.2 | USA | 29.8 | USA | 43.8 |

**Males**

| 20-39 | | 40-54 | | 55-69 | | 70-85+ | |
|---|---|---|---|---|---|---|---|
| UAE | 0.10 | UAE | 0.74 | Mauritania | 3.3 | Mauritania | |
| Somalia | 0.11 | Mauritania | 1.20 | Somalia | 4.1 | Sudan | 4.4 |
| Qatar | 0.15 | Somalia | 1.50 | Sudan | 4.9 | Somalia | 5.0 |
| Bahrain | 0.17 | Algeria | 1.80 | Algeria | 6.0 | Bahrain | 8.0 |
| Morocco | 0.18 | Qatar | 1.90 | Morocco | 7.0 | Oman | 8.1 |
| Sudan | 0.22 | Sudan | 2.20 | Oman | 7.5 | Algeria | 11.6 |
| Kuwait | 0.23 | Morocco | 2.40 | Bahrain | 8.0 | Iraq | 12.0 |
| Algeria | 0.28 | Kuwait | 2.70 | Qatar | 8.5 | Morocco | 14.5 |
| Mauritania | 0.29 | Tunisia | 3.10 | Tunisia | 8.8 | Tunisia | 18.5 |
| Libya | 0.30 | Bahrain | 3.60 | Iraq | 9.2 | Egypt | 19.3 |
| Tunisia | 0.34 | Oman | 4.00 | Kuwait | 9.9 | S.Arabia | 20.9 |
| Egypt | 0.46 | Egypt | 4.10 | Egypt | 11.0 | Syria | 22.0 |
| Iraq | 0.50 | Iraq | 4.50 | S.Arabia | 12.1 | Qatar | 22.1 |
| Jordan | 0.67 | Libya | 4.60 | Jordan | 13.5 | Kuwait | 23.3 |
| S.Arabia | 0.67 | S.Arabia | 4.80 | Palestine | 13.9 | Jordan | 23.6 |
| **Worldwide** | 0.71 | Jordan | 5.80 | Libya | 14.1 | Lebanon | 29.7 |
| Oman | 0.73 | Palestine | 7.00 | UAE | 14.5 | UAE | 36.8 |
| Syria | 0.98 | **Worldwide** | 7.20 | Syria | 14.6 | **Worldwide** | 42.6 |
| Palestine | 1.10 | Syria | 7.20 | Lebanon | 22.0 | Libya | 44.2 |
| Lebanon | 1.30 | Lebanon | 11.40 | **Worldwide** | 24.0 | Palestine | 48.6 |
| Europe | 1.80 | Europe | 18.70 | Europe | 55.3 | Europe | 80.0 |
| USA | 2.40 | USA | 20.70 | USA | 58.7 | USA | 85.9 |

### Larynx

#### Females

| 30-54 | | 55-69 | | 70-85+ | |
|---|---|---|---|---|---|
| Qatar | | Qatar | | Qatar | |
| Bahrain | | Bahrain | | Bahrain | |
| Kuwait | | Sudan | | UAE | |
| Mauritania | | S.Arabia | 0.82 | Mauritania | |
| UAE | | Oman | 0.98 | Somalia | 0.83 |
| Sudan | 0.05 | Jordan | 1.10 | S.Arabia | 0.99 |
| S.Arabia | 0.10 | Algeria | 1.20 | Tunisia | 1.90 |
| Morocco | 0.18 | Egypt | 1.20 | Egypt | 2.00 |
| Oman | 0.20 | Tunisia | 1.20 | Algeria | 2.30 |
| Algeria | 0.30 | Palestine | 1.50 | Europe | 2.70 |
| Somalia | 0.34 | Morocco | 1.60 | **Worldwide** | 2.90 |
| Egypt | 0.36 | Somalia | 1.70 | Oman | 3.20 |
| Libya | 0.38 | UAE | 2.00 | Libya | 3.30 |
| Tunisia | 0.39 | **Worldwide** | 2.10 | Jordan | 3.60 |
| **Worldwide** | 0.40 | Mauritania | 2.20 | Morocco | 3.60 |
| Palestine | 0.43 | Kuwait | 2.40 | Sudan | 3.80 |
| Europe | 0.62 | Europe | 3.00 | Iraq | 4.00 |
| Jordan | 0.69 | Libya | 3.60 | Syria | 5.40 |
| Iraq | 0.87 | Syria | 3.90 | USA | 5.50 |
| USA | 0.99 | Lebanon | 4.10 | Palestine | 6.80 |
| Syria | 1.20 | USA | 4.80 | Lebanon | 7.70 |
| Lebanon | 2.30 | Iraq | 6.20 | Kuwait | 8.10 |

#### Males

| 30-54 | | 55-69 | | 70-85+ | |
|---|---|---|---|---|---|
| UAE | 0.33 | S.Arabia | 4.5 | S.Arabia | 7.3 |
| S.Arabia | 0.63 | Bahrain | 4.5 | Oman | 8.1 |
| Kuwait | 0.67 | Somalia | 5.3 | Mauritania | 8.2 |
| Somalia | 0.73 | Mauritania | 6.7 | Somalia | 10.3 |
| Qatar | 0.87 | Oman | 6.7 | Qatar | 10.7 |
| Bahrain | 0.91 | Kuwait | 7.2 | UAE | 11.2 |
| Mauritania | 1.20 | Qatar | 7.8 | Sudan | 13.5 |
| Egypt | 1.40 | Sudan | 8.0 | Kuwait | 15.4 |
| Sudan | 1.70 | UAE | 8.5 | Egypt | 23.4 |
| Iraq | 1.90 | Egypt | 9.9 | **Worldwide** | 23.5 |
| Oman | 2.10 | **Worldwide** | 17.2 | Jordan | 26.0 |
| Algeria | 2.70 | Algeria | 19.2 | Europe | 27.8 |
| **Worldwide** | 2.70 | Palestine | 20.2 | USA | 30.1 |
| Libya | 3.10 | Jordan | 20.2 | Algeria | 30.6 |
| Tunisia | 3.30 | Iraq | 20.7 | Iraq | 31.9 |
| USA | 3.30 | USA | 21.1 | Morocco | 34.7 |
| Europe | 4.40 | Syria | 23.3 | Bahrain | 35.0 |
| Morocco | 4.60 | Libya | 25.1 | Syria | 36.2 |
| Syria | 4.60 | Tunisia | 26.1 | Tunisia | 43.2 |
| Palestine | 5.00 | Morocco | 28.4 | Lebanon | 49.1 |
| Jordan | 5.80 | Europe | 28.6 | Palestine | 53.4 |
| Lebanon | 5.80 | Lebanon | 30.1 | Libya | 53.6 |

### Leukaemia

**Females**

| 0-14 | | 15-34 | | 35-54 | | 55-69 | | 70-85+ | |
|---|---|---|---|---|---|---|---|---|---|
| Mauritania | 0.22 | Mauritania | 0.12 | Mauritania | 1.0 | Mauritania | 3.0 | Mauritania | |
| Morocco | 1.40 | Bahrain | 0.97 | Oman | 2.7 | Tunisia | 5.6 | Oman | 8.7 |
| Somalia | 1.70 | UAE | 1.10 | Qatar | 2.8 | Morocco | 6.6 | Algeria | 9.8 |
| Sudan | 1.90 | Morocco | 1.30 | Morocco | 3.0 | Oman | 6.7 | Morocco | 13.5 |
| Bahrain | 2.10 | Lebanon | 1.50 | Bahrain | 3.1 | Algeria | 7.3 | Iraq | 15.1 |
| Algeria | 2.40 | Algeria | 1.70 | Tunisia | 3.1 | Qatar | 9.0 | Qatar | 15.2 |
| Egypt | 2.40 | Jordan | 1.70 | Algeria | 3.3 | Bahrain | 9.5 | Libya | 15.2 |
| Jordan | 2.60 | Kuwait | 1.70 | Kuwait | 3.6 | Iraq | 9.8 | S.Arabia | 15.4 |
| Palestine | 2.60 | Somalia | 1.80 | **Worldwide** | 3.9 | **Worldwide** | 10.1 | UAE | 15.7 |
| **Worldwide** | 2.90 | Egypt | 1.80 | UAE | 4.0 | S.Arabia | 10.3 | Kuwait | 15.9 |
| Qatar | 2.90 | **Worldwide** | 1.80 | Palestine | 4.1 | Palestine | 11.1 | Tunisia | 16.9 |
| Tunisia | 3.10 | Syria | 1.80 | Jordan | 4.7 | Egypt | 12.0 | Egypt | 17.9 |
| Syria | 3.40 | Europe | 1.80 | Egypt | 4.7 | Lebanon | 12.3 | Sudan | 19.7 |
| Oman | 3.50 | Qatar | 1.90 | Europe | 4.7 | Syria | 12.9 | **Worldwide** | 21.2 |
| Libya | 3.60 | Sudan | 2.00 | Libya | 4.7 | Kuwait | 13.4 | Syria | 25.3 |
| Iraq | 3.70 | Palestine | 2.00 | S.Arabia | 4.9 | Sudan | 13.8 | Lebanon | 26.4 |
| Lebanon | 4.10 | Tunisia | 2.00 | Iraq | 5.0 | Somalia | 15.2 | Bahrain | 26.7 |
| UAE | 4.30 | Iraq | 2.10 | Syria | 5.1 | Europe | 15.4 | Somalia | 26.9 |
| Europe | 4.60 | Oman | 2.40 | Sudan | 5.7 | UAE | 15.7 | Europe | 32.2 |
| S.Arabia | 4.70 | S.Arabia | 2.40 | Lebanon | 6.1 | Jordan | 16.1 | Jordan | 34.2 |
| Kuwait | 5.60 | USA | 2.60 | Somalia | 6.3 | Libya | 17.2 | USA | 42.8 |
| USA | 5.70 | Libya | 2.90 | USA | 6.6 | USA | 20.7 | Palestine | 60.0 |

**Males**

| 0-14 | | 15-34 | | 35-54 | | 55-69 | | 70-85+ | |
|---|---|---|---|---|---|---|---|---|---|
| Mauritania | 0.54 | Mauritania | 0.37 | UAE | 0.84 | Mauritania | 2.5 | Mauritania | 2.9 |
| Sudan | 2.40 | Jordan | 1.50 | Mauritania | 0.99 | Morocco | 7.5 | UAE | 12.1 |
| Jordan | 2.50 | Sudan | 1.80 | Bahrain | 2.80 | UAE | 10.0 | Somalia | 17.8 |
| Somalia | 2.80 | Libya | 1.80 | Morocco | 3.00 | Tunisia | 10.2 | Tunisia | 17.9 |
| Morocco | 2.90 | Kuwait | 1.80 | Kuwait | 3.20 | Algeria | 10.6 | Oman | 20.5 |
| Libya | 3.00 | UAE | 1.80 | Libya | 3.30 | S.Arabia | 11.5 | Egypt | 21.0 |
| Tunisia | 3.20 | Morocco | 2.00 | Algeria | 3.40 | Bahrain | 11.9 | Qatar | 22.1 |
| Egypt | 3.30 | Algeria | 2.00 | S.Arabia | 3.80 | Palestine | 12.3 | Algeria | 23.0 |
| Algeria | 3.50 | Qatar | 2.00 | Jordan | 3.90 | Qatar | 13.1 | S.Arabia | 24.8 |
| Palestine | 3.50 | Palestine | 2.20 | Qatar | 4.00 | Oman | 13.1 | Morocco | 27.2 |
| **Worldwide** | 3.80 | Oman | 2.20 | **Worldwide** | 4.60 | Somalia | 13.5 | Iraq | 32.3 |
| Iraq | 3.80 | Europe | 2.20 | Tunisia | 4.80 | Egypt | 13.8 | Sudan | 34.2 |
| Syria | 3.80 | Bahrain | 2.30 | Syria | 4.90 | Kuwait | 14.8 | **Worldwide** | 36.5 |
| Qatar | 4.20 | Egypt | 2.40 | Egypt | 5.00 | Iraq | 14.8 | Lebanon | 37.9 |
| Oman | 4.80 | **Worldwide** | 2.40 | Iraq | 5.20 | **Worldwide** | 15.2 | Bahrain | 39.9 |
| Kuwait | 5.10 | Syria | 2.40 | Somalia | 5.30 | Libya | 16.1 | Libya | 47.6 |
| S.Arabia | 5.10 | Somalia | 2.50 | Palestine | 5.50 | Lebanon | 17.9 | Kuwait | 52.1 |
| Lebanon | 5.20 | Tunisia | 2.60 | Europe | 6.00 | Syria | 20.8 | Syria | 60.2 |
| Europe | 5.30 | Iraq | 2.60 | Oman | 6.30 | Sudan | 24.3 | Europe | 60.6 |
| USA | 5.80 | Lebanon | 2.60 | Lebanon | 7.20 | Europe | 26.1 | USA | 78.2 |
| Bahrain | 6.10 | S.Arabia | 2.70 | Sudan | 8.90 | Jordan | 26.8 | Jordan | 112.6 |
| UAE | 9.10 | USA | 3.20 | USA | 9.10 | USA | 34.7 | Palestine | 138.4 |

#### Lip & Oral

**Females**

| 20-39 | | 40-54 | | 55-69 | | 70-85+ | |
|---|---|---|---|---|---|---|---|
| Qatar | | UAE | 0.39 | UAE | 1.5 | UAE | |
| Bahrain | | Palestine | 0.67 | Algeria | 1.6 | Bahrain | |
| Libya | | Algeria | 0.75 | Tunisia | 3.3 | Algeria | 3.4 |
| UAE | 0.06 | Morocco | 0.85 | Lebanon | 3.5 | Jordan | 5.5 |
| Oman | 0.12 | Bahrain | 0.98 | Libya | 3.5 | Oman | 5.8 |
| Palestine | 0.15 | Qatar | 1.10 | Syria | 3.6 | Iraq | 6.9 |
| Algeria | 0.20 | Iraq | 1.10 | Palestine | 4.1 | Syria | 8.1 |
| Iraq | 0.21 | Jordan | 1.10 | Iraq | 4.1 | Libya | 8.4 |
| Morocco | 0.21 | Syria | 1.30 | Jordan | 4.1 | Mauritania | 9.4 |
| Kuwait | 0.22 | Kuwait | 1.50 | Egypt | 4.1 | Sudan | 10.9 |
| Mauritania | 0.30 | Tunisia | 1.60 | Qatar | 4.4 | Lebanon | 11.1 |
| S.Arabia | 0.31 | Egypt | 1.60 | Oman | 4.5 | S.Arabia | 11.3 |
| Syria | 0.31 | S.Arabia | 1.90 | Morocco | 5.1 | Tunisia | 11.5 |
| Jordan | 0.32 | Lebanon | 1.90 | S.Arabia | 5.1 | Morocco | 12.3 |
| Tunisia | 0.32 | Sudan | 2.00 | Mauritania | 5.3 | Somalia | 12.9 |
| Lebanon | 0.34 | Libya | 2.20 | Kuwait | 5.8 | Egypt | 14.3 |
| Somalia | 0.38 | Mauritania | 2.20 | Sudan | 7.5 | **Worldwide** | 14.4 |
| Egypt | 0.40 | Somalia | 2.40 | Bahrain | 8.0 | Europe | 14.9 |
| Europe | 0.44 | **Worldwide** | 3.40 | Somalia | 8.2 | Palestine | 16.1 |
| Sudan | 0.47 | USA | 3.50 | **Worldwide** | 8.4 | USA | 16.3 |
| **Worldwide** | 0.64 | Europe | 3.60 | Europe | 9.0 | Kuwait | 24.4 |
| USA | 0.66 | Oman | 4.00 | USA | 9.1 | Qatar | 28.0 |

**Males**

| 20-39 | | 40-54 | | 55-69 | | 70-85+ | |
|---|---|---|---|---|---|---|---|
| UAE | 0.02 | UAE | 0.26 | UAE | 2.6 | Bahrain | |
| Libya | 0.10 | Iraq | 0.68 | Algeria | 3.3 | UAE | 4.0 |
| Tunisia | 0.24 | Algeria | 1.30 | Mauritania | 3.3 | Oman | 4.0 |
| Morocco | 0.25 | Libya | 1.40 | Palestine | 4.7 | Algeria | 6.1 |
| Iraq | 0.27 | S.Arabia | 1.40 | Lebanon | 4.9 | Libya | 8.6 |
| Bahrain | 0.29 | Bahrain | 1.60 | Libya | 5.4 | Iraq | 9.6 |
| Lebanon | 0.31 | Syria | 1.80 | Jordan | 5.5 | Somalia | 10.8 |
| Algeria | 0.32 | Mauritania | 1.90 | Syria | 5.8 | Syria | 11.4 |
| Egypt | 0.32 | Kuwait | 1.90 | S.Arabia | 6.0 | Jordan | 12.1 |
| Qatar | 0.32 | Qatar | 2.20 | Iraq | 6.6 | S.Arabia | 12.8 |
| S.Arabia | 0.32 | Lebanon | 2.30 | Kuwait | 6.8 | Lebanon | 13.0 |
| Syria | 0.32 | Palestine | 2.40 | Tunisia | 7.5 | Sudan | 13.3 |
| Jordan | 0.37 | Tunisia | 2.60 | Qatar | 7.7 | Egypt | 13.4 |
| Sudan | 0.37 | Egypt | 2.60 | Egypt | 7.7 | Morocco | 13.9 |
| Somalia | 0.38 | Jordan | 2.60 | Somalia | 9.2 | Mauritania | 14.6 |
| Palestine | 0.40 | Morocco | 3.10 | Sudan | 9.3 | Qatar | 15.5 |
| Mauritania | 0.44 | Somalia | 3.10 | Morocco | 9.7 | Tunisia | 18.7 |
| Kuwait | 0.45 | Sudan | 3.80 | Oman | 11.5 | Kuwait | 18.9 |
| Oman | 0.49 | Oman | 4.50 | Bahrain | 12.4 | Palestine | 24.2 |
| Europe | 0.77 | USA | 9.10 | **Worldwide** | 21.8 | **Worldwide** | 28.8 |
| USA | 0.79 | **Worldwide** | 9.60 | USA | 28.9 | Europe | 32.4 |
| **Worldwide** | 2.10 | Europe | 10.90 | Europe | 30.9 | USA | 34.2 |

### Liver

#### Females

| 30-54 | |
|---|---|
| Palestine | 0.22 |
| Morocco | 0.46 |
| Bahrain | 0.58 |
| Qatar | 0.63 |
| Algeria | 1.00 |
| Kuwait | 1.10 |
| Iraq | 1.30 |
| Jordan | 1.30 |
| S.Arabia | 1.40 |
| Europe | 1.50 |
| Syria | 1.50 |
| UAE | 1.50 |
| Libya | 1.70 |
| Lebanon | 1.90 |
| USA | 2.40 |
| Oman | 2.70 |
| Sudan | 2.90 |
| Tunisia | 3.30 |
| **Worldwide** | 3.70 |
| Somalia | 3.70 |
| Mauritania | 9.80 |
| Egypt | 10.90 |

| 55-69 | |
|---|---|
| Morocco | 4.1 |
| Palestine | 4.9 |
| Algeria | 5.5 |
| Jordan | 8.4 |
| Tunisia | 8.8 |
| Syria | 9.2 |
| Iraq | 9.8 |
| Europe | 9.8 |
| Lebanon | 10.1 |
| S.Arabia | 10.6 |
| Sudan | 10.7 |
| Oman | 11.2 |
| UAE | 13.0 |
| Bahrain | 14.5 |
| Qatar | 14.6 |
| USA | 14.9 |
| Somalia | 14.9 |
| Libya | 15.1 |
| Kuwait | 16.6 |
| **Worldwide** | 19.1 |
| Mauritania | 21.9 |
| Egypt | 74.7 |

| 70-85+ | |
|---|---|
| Qatar | |
| Algeria | 8.1 |
| Morocco | 8.9 |
| Somalia | 11.6 |
| Tunisia | 14.3 |
| Palestine | 15.5 |
| Oman | 15.9 |
| Sudan | 18.4 |
| Iraq | 21.5 |
| S.Arabia | 21.8 |
| USA | 23.0 |
| Lebanon | 23.7 |
| Bahrain | 24.4 |
| Europe | 26.3 |
| Syria | 27.1 |
| Mauritania | 27.3 |
| Jordan | 33.5 |
| Kuwait | 36.0 |
| Libya | 38.9 |
| **Worldwide** | 41.5 |
| UAE | 58.2 |
| Egypt | 128.6 |

#### Males

| 30-54 | |
|---|---|
| UAE | 0.30 |
| Morocco | 0.66 |
| Algeria | 0.71 |
| Iraq | 1.10 |
| S.Arabia | 1.10 |
| Jordan | 1.60 |
| Syria | 1.60 |
| Palestine | 1.70 |
| Qatar | 1.70 |
| Bahrain | 1.80 |
| Kuwait | 1.90 |
| Lebanon | 2.40 |
| Tunisia | 2.70 |
| Libya | 2.80 |
| Somalia | 4.10 |
| Oman | 4.20 |
| Europe | 4.90 |
| Sudan | 5.50 |
| USA | 9.10 |
| **Worldwide** | 13.80 |
| Mauritania | 18.70 |
| Egypt | 45.70 |

| 55-69 | |
|---|---|
| Morocco | 5.2 |
| Algeria | 6.8 |
| Palestine | 8.3 |
| Tunisia | 9.7 |
| UAE | 12.3 |
| Jordan | 13.0 |
| Somalia | 13.2 |
| Iraq | 14.1 |
| Syria | 14.3 |
| Lebanon | 14.6 |
| Bahrain | 16.0 |
| Libya | 17.1 |
| Sudan | 19.0 |
| Qatar | 19.3 |
| S.Arabia | 22.1 |
| Kuwait | 24.0 |
| Oman | 25.4 |
| Europe | 34.6 |
| USA | 48.6 |
| **Worldwide** | 55.4 |
| Mauritania | 63.6 |
| Egypt | 216.7 |

| 70-85+ | |
|---|---|
| Morocco | 13.8 |
| Tunisia | 14.5 |
| Algeria | 14.6 |
| Somalia | 22.2 |
| Palestine | 24.3 |
| Sudan | 24.6 |
| Iraq | 28.5 |
| Syria | 32.9 |
| Lebanon | 32.9 |
| Jordan | 35.7 |
| Bahrain | 39.9 |
| Libya | 40.0 |
| Oman | 40.9 |
| USA | 54.0 |
| Europe | 66.2 |
| UAE | 69.9 |
| Kuwait | 75.8 |
| Qatar | 78.4 |
| S.Arabia | 83.1 |
| **Worldwide** | 90.4 |
| Mauritania | 94.3 |
| Egypt | 295.5 |

### Lung

#### Females

| 20-54 | | 55-69 | | 70-85+ | |
|---|---|---|---|---|---|
| Mauritania | 0.70 | Mauritania | 5.9 | Mauritania | 7.7 |
| S.Arabia | 0.82 | Sudan | 6.5 | Oman | 7.8 |
| UAE | 0.96 | Oman | 6.7 | Sudan | 8.3 |
| Qatar | 1.00 | Tunisia | 9.7 | Tunisia | 14.0 |
| Oman | 1.10 | S.Arabia | 11.0 | Algeria | 18.6 |
| Libya | 1.40 | Qatar | 12.2 | Libya | 19.0 |
| Sudan | 1.40 | Algeria | 12.7 | Somalia | 19.5 |
| Kuwait | 1.50 | Somalia | 13.0 | Morocco | 21.6 |
| Algeria | 1.60 | Libya | 14.1 | S.Arabia | 23.5 |
| Somalia | 1.90 | Morocco | 14.6 | Egypt | 32.3 |
| Tunisia | 2.00 | Egypt | 15.8 | Iraq | 36.8 |
| Bahrain | 2.20 | Palestine | 18.9 | UAE | 41.9 |
| Iraq | 2.20 | Kuwait | 19.0 | Palestine | 45.9 |
| Morocco | 2.20 | Bahrain | 20.8 | Jordan | 53.5 |
| Palestine | 2.40 | Iraq | 22.7 | Kuwait | 62.4 |
| Jordan | 2.50 | Jordan | 24.3 | Syria | 66.2 |
| Egypt | 2.60 | UAE | 26.7 | Bahrain | 84.7 |
| Syria | 4.50 | Syria | 35.3 | Qatar | 100.2 |
| **Worldwide** | 5.40 | Lebanon | 59.3 | Lebanon | 107.5 |
| Europe | 8.00 | **Worldwide** | 61.1 | Europe | 129.8 |
| Lebanon | 8.80 | Europe | 86.2 | **Worldwide** | 134.6 |
| USA | 10.50 | USA | 132.9 | USA | 286.7 |

#### Males

| 20-54 | | 55-69 | | 70-85+ | |
|---|---|---|---|---|---|
| UAE | 0.38 | Sudan | 11.9 | Sudan | 19.5 |
| Kuwait | 0.89 | Somalia | 15.8 | Somalia | 27.2 |
| Qatar | 1.10 | Mauritania | 16.2 | Mauritania | 33.7 |
| Bahrain | 1.40 | S.Arabia | 19.7 | Oman | 51.1 |
| S.Arabia | 1.50 | Qatar | 33.7 | UAE | 68.5 |
| Mauritania | 1.70 | Kuwait | 34.5 | S.Arabia | 72.8 |
| Sudan | 1.80 | Oman | 36.3 | Egypt | 95.6 |
| Somalia | 1.90 | UAE | 36.4 | Algeria | 109.5 |
| Oman | 2.30 | Bahrain | 42.0 | Kuwait | 112.1 |
| Iraq | 5.20 | Egypt | 47.7 | Iraq | 127.8 |
| Egypt | 5.30 | Algeria | 87.6 | Qatar | 151.2 |
| Algeria | 7.30 | Iraq | 90.6 | Tunisia | 165.3 |
| Palestine | 9.00 | Jordan | 106.5 | Morocco | 166.2 |
| **Worldwide** | 9.50 | Palestine | 107.4 | Libya | 217.8 |
| Syria | 9.80 | Syria | 111.9 | Syria | 247.0 |
| USA | 10.40 | Libya | 116.7 | Lebanon | 256.8 |
| Libya | 10.70 | Tunisia | 117.2 | Palestine | 271.1 |
| Jordan | 12.20 | Lebanon | 134.5 | Bahrain | 277.1 |
| Europe | 13.20 | **Worldwide** | 137.8 | **Worldwide** | 301.7 |
| Lebanon | 13.80 | Morocco | 157.4 | Europe | 364.3 |
| Tunisia | 15.40 | USA | 172.3 | Jordan | 368.4 |
| Morocco | 17.60 | Europe | 215.7 | USA | 412.7 |

### Melanoma

#### Females

### 20-54

| | |
|---|---|
| Qatar | |
| Bahrain | |
| S.Arabia | 0.17 |
| Egypt | 0.19 |
| Libya | 0.24 |
| Kuwait | 0.27 |
| Jordan | 0.30 |
| Morocco | 0.31 |
| Sudan | 0.32 |
| Somalia | 0.35 |
| Iraq | 0.38 |
| Mauritania | 0.41 |
| Algeria | 0.44 |
| UAE | 0.48 |
| Tunisia | 0.50 |
| Palestine | 0.61 |
| Oman | 0.65 |
| Syria | 0.65 |
| Lebanon | 1.20 |
| **Worldwide** | 2.50 |
| USA | 11.80 |
| Europe | 14.00 |

### 55-69

| | |
|---|---|
| Qatar | |
| Bahrain | |
| UAE | |
| S.Arabia | 0.94 |
| Oman | 0.98 |
| Tunisia | 1.20 |
| Egypt | 1.40 |
| Kuwait | 1.50 |
| Sudan | 1.60 |
| Jordan | 1.80 |
| Algeria | 1.80 |
| Iraq | 1.90 |
| Libya | 2.30 |
| Syria | 2.50 |
| Morocco | 2.90 |
| Somalia | 3.00 |
| Palestine | 3.20 |
| Lebanon | 4.10 |
| Mauritania | 4.70 |
| **Worldwide** | 9.20 |
| Europe | 28.90 |
| USA | 33.10 |

### 70-85+

| | |
|---|---|
| Qatar | |
| Bahrain | |
| UAE | |
| Kuwait | |
| Libya | |
| Iraq | 0.66 |
| S.Arabia | 1.30 |
| Egypt | 1.70 |
| Jordan | 2.70 |
| Oman | 3.20 |
| Tunisia | 3.60 |
| Somalia | 3.60 |
| Palestine | 3.70 |
| Morocco | 4.40 |
| Sudan | 4.60 |
| Algeria | 5.30 |
| Syria | 5.60 |
| Mauritania | 5.90 |
| Lebanon | 13.30 |
| **Worldwide** | 17.70 |
| Europe | 39.40 |
| USA | 48.20 |

#### Males

### 20-54

| | |
|---|---|
| Mauritania | |
| Libya | |
| Bahrain | |
| UAE | 0.02 |
| S.Arabia | 0.02 |
| Egypt | 0.03 |
| Morocco | 0.10 |
| Sudan | 0.11 |
| Qatar | 0.11 |
| Iraq | 0.12 |
| Oman | 0.13 |
| Jordan | 0.15 |
| Algeria | 0.16 |
| Tunisia | 0.17 |
| Somalia | 0.18 |
| Kuwait | 0.21 |
| Syria | 0.22 |
| **Worldwide** | 0.34 |
| Palestine | 0.36 |
| Lebanon | 0.39 |
| USA | 0.89 |
| Europe | 1.40 |

### 55-69

| | |
|---|---|
| Qatar | |
| S.Arabia | 0.44 |
| Egypt | 0.81 |
| Libya | 1.10 |
| Jordan | 1.10 |
| Bahrain | 1.50 |
| Palestine | 1.50 |
| Iraq | 1.60 |
| Kuwait | 1.60 |
| Mauritania | 1.80 |
| Tunisia | 1.90 |
| Somalia | 2.00 |
| Algeria | 2.10 |
| UAE | 2.20 |
| Morocco | 2.40 |
| Sudan | 2.70 |
| Oman | 3.10 |
| Syria | 3.10 |
| Lebanon | 6.20 |
| **Worldwide** | 12.20 |
| Europe | 37.40 |
| USA | 55.50 |

### 70-85+

| | |
|---|---|
| Qatar | |
| Bahrain | |
| Iraq | 0.84 |
| Sudan | 2.20 |
| Jordan | 2.40 |
| Somalia | 2.50 |
| Kuwait | 2.70 |
| Mauritania | 2.90 |
| S.Arabia | 3.30 |
| Libya | 3.60 |
| Egypt | 3.90 |
| Oman | 4.10 |
| Tunisia | 4.40 |
| Morocco | 5.60 |
| Palestine | 6.30 |
| UAE | 6.30 |
| Algeria | 6.80 |
| Syria | 7.20 |
| Lebanon | 17.30 |
| **Worldwide** | 30.80 |
| Europe | 67.60 |
| USA | 123.60 |

#### Mesothelioma

##### Females

**25-54**

| Country | Value |
|---|---|
| Qatar | |
| Oman | |
| Bahrain | |
| Libya | |
| Palestine | |
| Mauritania | |
| Morocco | 0.04 |
| Sudan | 0.04 |
| Jordan | 0.07 |
| Somalia | 0.07 |
| Algeria | 0.08 |
| Tunisia | 0.08 |
| S.Arabia | 0.09 |
| **Worldwide** | 0.11 |
| Kuwait | 0.13 |
| UAE | 0.15 |
| USA | 0.15 |
| Europe | 0.16 |
| Iraq | 0.16 |
| Syria | 0.19 |
| Egypt | 0.20 |
| Lebanon | 0.39 |

**55-69**

| Country | Value |
|---|---|
| Qatar | |
| Oman | |
| Bahrain | |
| Libya | |
| Palestine | |
| Mauritania | |
| Jordan | |
| Kuwait | |
| UAE | |
| Tunisia | 0.14 |
| Sudan | 0.20 |
| Algeria | 0.20 |
| S.Arabia | 0.26 |
| Syria | 0.39 |
| Somalia | 0.47 |
| Morocco | 0.58 |
| Lebanon | 0.60 |
| **Worldwide** | 0.65 |
| Iraq | 0.66 |
| Egypt | 0.68 |
| USA | 0.90 |
| Europe | 1.20 |

**70-85+**

| Country | Value |
|---|---|
| Qatar | |
| Oman | |
| Bahrain | |
| Libya | |
| Palestine | |
| Mauritania | |
| Jordan | |
| Kuwait | |
| UAE | |
| Tunisia | |
| Sudan | |
| S.Arabia | |
| Syria | |
| Somalia | |
| Lebanon | |
| Iraq | |
| Algeria | 0.50 |
| Morocco | 0.98 |
| **Worldwide** | 1.60 |
| USA | 2.40 |
| Egypt | 2.80 |
| Europe | 3.20 |

##### Males

**25-54**

| Country | Value |
|---|---|
| Bahrain | |
| Palestine | |
| Somalia | |
| Sudan | |
| UAE | |
| Mauritania | |
| Tunisia | 0.04 |
| Oman | 0.06 |
| Kuwait | 0.08 |
| Qatar | 0.08 |
| S.Arabia | 0.12 |
| **Worldwide** | 0.13 |
| Iraq | 0.13 |
| Jordan | 0.13 |
| Algeria | 0.14 |
| Egypt | 0.15 |
| Morocco | 0.20 |
| USA | 0.21 |
| Syria | 0.24 |
| Libya | 0.26 |
| Europe | 0.28 |
| Lebanon | 0.50 |

**55-69**

| Country | Value |
|---|---|
| Sudan | |
| Mauritania | |
| Somalia | 0.30 |
| Tunisia | 0.46 |
| S.Arabia | 0.59 |
| Morocco | 0.72 |
| UAE | 0.81 |
| Egypt | 0.87 |
| Algeria | 1.00 |
| Iraq | 1.10 |
| Libya | 1.10 |
| Jordan | 1.30 |
| Palestine | 1.50 |
| **Worldwide** | 1.50 |
| Oman | 1.60 |
| Syria | 2.00 |
| USA | 3.30 |
| Lebanon | 3.80 |
| Europe | 4.00 |
| Qatar | 5.10 |
| Kuwait | 5.50 |
| Bahrain | 7.40 |

**70-85+**

| Country | Value |
|---|---|
| Qatar | |
| Oman | |
| Sudan | |
| Mauritania | |
| UAE | |
| Iraq | 1.4 |
| Somalia | 1.6 |
| Jordan | 1.7 |
| Tunisia | 1.8 |
| Algeria | 1.9 |
| Morocco | 2.0 |
| Syria | 2.1 |
| S.Arabia | 2.6 |
| Libya | 3.6 |
| Lebanon | 3.6 |
| Egypt | 4.3 |
| Palestine | 4.3 |
| **Worldwide** | 6.7 |
| Kuwait | 9.2 |
| Bahrain | 9.6 |
| USA | 13.5 |
| Europe | 17.3 |

### Multiple Myeloma

#### Females

| 20-54 | | 55-69 | | 70-85+ | |
|---|---|---|---|---|---|
| Qatar | | S.Arabia | 4.8 | Qatar | |
| Mauritania | | Morocco | 5.1 | Mauritania | |
| UAE | 0.28 | Jordan | 5.2 | Sudan | 1.7 |
| Iraq | 0.30 | Palestine | 5.3 | Somalia | 3.0 |
| Sudan | 0.49 | Oman | 5.7 | Egypt | 4.0 |
| Egypt | 0.50 | Sudan | 5.9 | Morocco | 4.0 |
| S.Arabia | 0.51 | Tunisia | 6.6 | Iraq | 4.1 |
| Algeria | 0.53 | Somalia | 7.3 | Oman | 5.5 |
| Kuwait | 0.56 | Mauritania | 7.5 | Libya | 6.6 |
| **Worldwide** | 0.57 | Algeria | 7.8 | Tunisia | 6.7 |
| Tunisia | 0.58 | Bahrain | 8.0 | S.Arabia | 7.3 |
| Morocco | 0.68 | **Worldwide** | 8.7 | Bahrain | 7.4 |
| Libya | 0.74 | Kuwait | 9.3 | Syria | 11.4 |
| Oman | 0.77 | Libya | 10.0 | Kuwait | 12.2 |
| Somalia | 0.85 | Qatar | 12.2 | **Worldwide** | 12.7 |
| Europe | 0.89 | UAE | 14.6 | Jordan | 12.8 |
| Syria | 0.94 | Iraq | 16.3 | Algeria | 13.8 |
| Bahrain | 1.10 | Europe | 17.0 | Lebanon | 18.6 |
| Jordan | 1.20 | Syria | 19.2 | Europe | 20.9 |
| Lebanon | 1.30 | USA | 20.6 | USA | 29.1 |
| Palestine | 1.30 | Egypt | 24.0 | Palestine | 31.1 |
| USA | 1.40 | Lebanon | 37.0 | UAE | 34.9 |

#### Males

| 20-54 | | 55-69 | | 70-85 | |
|---|---|---|---|---|---|
| Mauritania | 0.15 | Mauritania | 0.97 | Mauritania | |
| UAE | 0.18 | UAE | 3.30 | Sudan | 3.2 |
| Bahrain | 0.24 | S.Arabia | 4.70 | Egypt | 3.3 |
| Libya | 0.28 | Qatar | 5.00 | Iraq | 6.9 |
| S.Arabia | 0.52 | Iraq | 5.00 | Bahrain | 8.0 |
| Somalia | 0.56 | Tunisia | 5.10 | Somalia | 8.3 |
| Qatar | 0.60 | Bahrain | 5.40 | Tunisia | 9.2 |
| Tunisia | 0.60 | Egypt | 6.00 | Kuwait | 9.2 |
| Kuwait | 0.64 | Morocco | 6.20 | S.Arabia | 9.6 |
| Iraq | 0.73 | Kuwait | 7.20 | Qatar | 10.7 |
| **Worldwide** | 0.76 | Somalia | 7.50 | UAE | 11.9 |
| Morocco | 0.80 | Algeria | 7.50 | Oman | 12.3 |
| Algeria | 0.84 | Libya | 8.00 | Algeria | 13.8 |
| Egypt | 1.20 | Oman | 8.20 | Morocco | 13.9 |
| Oman | 1.20 | **Worldwide** | 8.30 | Syria | 14.3 |
| Sudan | 1.30 | Syria | 8.60 | Libya | 15.9 |
| Europe | 1.40 | Sudan | 9.70 | Jordan | 17.0 |
| Syria | 1.40 | Lebanon | 9.90 | Lebanon | 19.6 |
| Jordan | 1.80 | Jordan | 10.20 | **Worldwide** | 20.1 |
| Lebanon | 1.80 | Palestine | 11.50 | Europe | 35.3 |
| USA | 1.90 | Europe | 14.20 | Palestine | 42.3 |
| Palestine | 2.30 | USA | 20.60 | USA | 46.5 |

### Nasopharynx

#### Females

### 20-39

| Country | Value |
|---|---|
| Qatar | |
| Mauritania | |
| UAE | |
| Bahrain | |
| Kuwait | |
| Egypt | 0.14 |
| Europe | 0.14 |
| USA | 0.14 |
| Oman | 0.15 |
| Palestine | 0.24 |
| Lebanon | 0.28 |
| Syria | 0.31 |
| Iraq | 0.35 |
| Jordan | 0.38 |
| S.Arabia | 0.38 |
| **Worldwide** | 0.53 |
| Somalia | 0.71 |
| Sudan | 0.94 |
| Libya | 0.96 |
| Tunisia | 1.10 |
| Algeria | 1.20 |
| Morocco | 1.20 |

### 40-69

| Country | Value |
|---|---|
| Qatar | |
| Mauritania | |
| UAE | 0.20 |
| Europe | 0.50 |
| Egypt | 0.52 |
| USA | 0.54 |
| Oman | 0.57 |
| Bahrain | 0.62 |
| Palestine | 0.65 |
| Iraq | 0.66 |
| Kuwait | 1.10 |
| Syria | 1.10 |
| Jordan | 1.10 |
| S.Arabia | 1.40 |
| Lebanon | 1.50 |
| **Worldwide** | 2.00 |
| Somalia | 2.60 |
| Sudan | 2.80 |
| Tunisia | 3.10 |
| Morocco | 3.40 |
| Libya | 3.50 |
| Algeria | 4.30 |

### 70-85+

| Country | Value |
|---|---|
| Qatar | |
| Mauritania | |
| UAE | |
| Oman | |
| Europe | 0.60 |
| Egypt | 0.93 |
| USA | 0.95 |
| Iraq | 0.97 |
| Syria | 1.40 |
| Jordan | 1.50 |
| S.Arabia | 1.60 |
| Lebanon | 1.70 |
| **Worldwide** | 2.10 |
| Libya | 2.50 |
| Somalia | 3.20 |
| Morocco | 3.20 |
| Tunisia | 3.30 |
| Kuwait | 4.10 |
| Sudan | 4.20 |
| Palestine | 4.60 |
| Algeria | 5.80 |
| Bahrain | 11.40 |

#### Males

### 20-39

| Country | Value |
|---|---|
| Qatar | |
| UAE | |
| Kuwait | 0.07 |
| Egypt | 0.14 |
| Oman | 0.23 |
| USA | 0.28 |
| Bahrain | 0.29 |
| Europe | 0.34 |
| Iraq | 0.36 |
| Lebanon | 0.39 |
| Mauritania | 0.45 |
| Syria | 0.48 |
| Libya | 0.61 |
| Sudan | 0.63 |
| Palestine | 0.64 |
| S.Arabia | 0.67 |
| Jordan | 0.69 |
| **Worldwide** | 1.10 |
| Somalia | 1.30 |
| Morocco | 1.50 |
| Tunisia | 1.70 |
| Algeria | 2.80 |

### 40-69

| Country | Value |
|---|---|
| UAE | 0.56 |
| Bahrain | 1.00 |
| Mauritania | 1.00 |
| Egypt | 1.50 |
| USA | 1.60 |
| Europe | 1.70 |
| Qatar | 1.80 |
| Oman | 2.40 |
| Lebanon | 2.40 |
| Iraq | 2.70 |
| Kuwait | 2.90 |
| Syria | 3.10 |
| Palestine | 3.10 |
| Jordan | 3.90 |
| Saudi Arabia | 4.10 |
| Sudan | 5.40 |
| Somalia | 5.50 |
| **Worldwide** | 5.80 |
| Libya | 7.10 |
| Tunisia | 7.40 |
| Morocco | 7.80 |
| Algeria | 11.30 |

### 70-85+

| Country | Value |
|---|---|
| Qatar | |
| Oman | |
| UAE | |
| Bahrain | |
| Mauritania | |
| Europe | 1.9 |
| USA | 2.1 |
| Lebanon | 2.8 |
| Somalia | 3.3 |
| Iraq | 3.4 |
| Palestine | 4.3 |
| Syria | 4.5 |
| S.Arabia | 4.9 |
| Sudan | 5.1 |
| Egypt | 5.6 |
| **Worldwide** | 5.9 |
| Morocco | 6.0 |
| Jordan | 6.1 |
| Kuwait | 7.1 |
| Libya | 9.5 |
| Tunisia | 11.6 |
| Algeria | 13.5 |

### non-Hodgkin's Lymphoma

#### Females

| 0-14 | | 10-24 | | 25-39 | | 40-69 | | 70-85+ | |
|---|---|---|---|---|---|---|---|---|---|
| UAE | | UAE | 0.19 | UAE | 0.87 | Mauritania | 3.3 | Mauritania | 9.4 |
| S.Arabia | 0.41 | Egypt | 0.44 | Mauritania | 1.10 | Algeria | 6.6 | Somalia | 9.8 |
| Kuwait | 0.45 | Sudan | 0.62 | Bahrain | 1.10 | Tunisia | 7.5 | Iraq | 18.0 |
| Jordan | 0.46 | Iraq | 0.64 | Libya | 1.40 | Sudan | 8.2 | Algeria | 21.0 |
| Morocco | 0.51 | Libya | 0.74 | S.Arabia | 1.50 | S.Arabia | 8.9 | Tunisia | 23.6 |
| Sudan | 0.51 | Oman | 0.75 | Morocco | 1.50 | Libya | 9.0 | Sudan | 24.7 |
| Oman | 0.59 | S.Arabia | 0.76 | Kuwait | 1.70 | Morocco | 9.1 | S.Arabia | 26.0 |
| Qatar | 0.59 | Jordan | 0.76 | Iraq | 1.80 | Somalia | 9.1 | Libya | 26.1 |
| **Worldwide** | 0.62 | Kuwait | 0.84 | Oman | 1.80 | Iraq | 9.4 | Qatar | 28.0 |
| Palestine | 0.62 | **Worldwide** | 0.84 | Tunisia | 1.90 | Qatar | 9.5 | Oman | 30.1 |
| Algeria | 0.63 | Morocco | 0.87 | Qatar | 1.90 | **Worldwide** | 9.6 | **Worldwide** | 32.6 |
| Somalia | 0.63 | Palestine | 0.99 | Sudan | 2.00 | Bahrain | 10.4 | Jordan | 33.0 |
| Syria | 0.67 | Mauritania | 0.99 | **Worldwide** | 2.00 | Palestine | 12.0 | Morocco | 36.4 |
| Tunisia | 0.67 | Somalia | 1.00 | Algeria | 2.40 | Jordan | 12.5 | Europe | 41.3 |
| Europe | 0.70 | Syria | 1.00 | Jordan | 2.50 | UAE | 12.8 | UAE | 41.9 |
| Bahrain | 0.73 | USA | 1.00 | Palestine | 2.90 | Oman | 14.1 | Bahrain | 48.0 |
| Iraq | 0.78 | Algeria | 1.10 | Europe | 2.90 | Europe | 14.1 | Kuwait | 50.2 |
| USA | 0.81 | Tunisia | 1.10 | Egypt | 3.30 | Kuwait | 14.5 | Syria | 50.9 |
| Lebanon | 0.84 | Europe | 1.20 | Syria | 3.50 | Syria | 14.6 | USA | 75.5 |
| Mauritania | 0.89 | Qatar | 1.40 | USA | 3.50 | Lebanon | 22.2 | Palestine | 79.2 |
| Egypt | 0.96 | Bahrain | 1.60 | Somalia | 4.10 | USA | 22.7 | Egypt | 82.8 |
| Libya | 1.20 | Lebanon | 2.00 | Lebanon | 6.10 | Egypt | 23.0 | Lebanon | 98.8 |

#### Males

| 0-14 | | 10-24 | | 25-39 | | 40-69 | | 70-85+ | |
|---|---|---|---|---|---|---|---|---|---|
| Oman | 0.39 | Qatar | 0.14 | UAE | 0.19 | Mauritania | 4.9 | Algeria | 19.3 |
| Jordan | 0.67 | UAE | 0.38 | Bahrain | 0.82 | UAE | 6.7 | Libya | 20.5 |
| Bahrain | 0.70 | Somalia | 0.80 | Qatar | 1.40 | Kuwait | 9.9 | Somalia | 23.1 |
| UAE | 0.75 | Oman | 0.84 | Mauritania | 1.50 | Qatar | 10.2 | Iraq | 23.2 |
| Palestine | 0.79 | Jordan | 0.95 | Kuwait | 2.20 | Algeria | 10.3 | Mauritania | 27.9 |
| Libya | 0.83 | Sudan | 1.00 | Libya | 2.30 | S.Arabia | 10.5 | Tunisia | 30.0 |
| Syria | 0.88 | Palestine | 1.10 | Iraq | 2.30 | Libya | 11.7 | Oman | 34.7 |
| Mauritania | 0.97 | Libya | 1.10 | **Worldwide** | 2.50 | Iraq | 11.7 | S.Arabia | 36.8 |
| Lebanon | 0.98 | Syria | 1.10 | Tunisia | 2.60 | Tunisia | 12.5 | Qatar | 39.3 |
| S.Arabia | 0.99 | Tunisia | 1.10 | S.Arabia | 2.60 | Bahrain | 12.7 | Jordan | 39.4 |
| Qatar | 1.00 | Bahrain | 1.20 | Algeria | 2.60 | **Worldwide** | 13.3 | Morocco | 44.3 |
| **Worldwide** | 1.10 | S.Arabia | 1.20 | Morocco | 3.10 | Somalia | 13.5 | **Worldwide** | 47.3 |
| Europe | 1.30 | **Worldwide** | 1.20 | Oman | 3.30 | Palestine | 13.6 | Egypt | 52.5 |
| USA | 1.30 | Algeria | 1.20 | Jordan | 3.30 | Morocco | 14.9 | Sudan | 53.5 |
| Algeria | 1.40 | Kuwait | 1.20 | Palestine | 3.40 | Jordan | 14.9 | UAE | 56.7 |
| Kuwait | 1.50 | Iraq | 1.20 | Somalia | 3.50 | Sudan | 16.6 | Syria | 59.1 |
| Tunisia | 1.50 | Egypt | 1.20 | Sudan | 3.70 | Oman | 17.5 | Europe | 66.1 |
| Morocco | 1.60 | Mauritania | 1.30 | Europe | 4.00 | Syria | 18.3 | Bahrain | 70.1 |
| Iraq | 1.70 | Europe | 1.60 | Syria | 4.20 | Europe | 20.4 | Kuwait | 76.1 |
| Somalia | 1.70 | USA | 1.60 | Egypt | 5.40 | Lebanon | 28.3 | Palestine | 99.9 |
| Sudan | 1.80 | Morocco | 1.70 | USA | 5.60 | Egypt | 30.5 | USA | 108.9 |
| Egypt | 2.00 | Lebanon | 2.00 | Lebanon | 6.90 | USA | 31.3 | Lebanon | 112.6 |

### Oesophagus

#### Females

| 20-39 | |
|---|---|
| Qatar | |
| Lebanon | |
| Palestine | |
| UAE | |
| Bahrain | |
| USA | 0.06 |
| Europe | 0.07 |
| Libya | 0.08 |
| Syria | 0.11 |
| Egypt | 0.12 |
| Iraq | 0.13 |
| Oman | 0.15 |
| S.Arabia | 0.16 |
| Tunisia | 0.18 |
| Jordan | 0.19 |
| Morocco | 0.23 |
| Kuwait | 0.24 |
| Mauritania | 0.30 |
| **Worldwide** | 0.31 |
| Somalia | 0.40 |
| Algeria | 0.43 |
| Sudan | 0.58 |

| 40-54 | |
|---|---|
| Qatar | |
| Kuwait | 0.27 |
| Libya | 0.38 |
| UAE | 0.57 |
| Iraq | 0.62 |
| Jordan | 0.73 |
| Palestine | 0.74 |
| Syria | 0.74 |
| S.Arabia | 0.78 |
| Egypt | 0.81 |
| Lebanon | 0.87 |
| Algeria | 0.94 |
| Bahrain | 0.98 |
| USA | 1.10 |
| Tunisia | 1.10 |
| Europe | 1.20 |
| Oman | 1.20 |
| Morocco | 1.30 |
| Mauritania | 1.50 |
| **Worldwide** | 3.30 |
| Sudan | 4.30 |
| Somalia | 8.90 |

| 55-69 | |
|---|---|
| Qatar | |
| Libya | |
| Lebanon | 1.9 |
| Tunisia | 2.0 |
| Palestine | 2.3 |
| Algeria | 2.3 |
| Syria | 2.5 |
| Kuwait | 2.6 |
| Jordan | 2.8 |
| Morocco | 3.2 |
| Iraq | 3.3 |
| S.Arabia | 3.4 |
| Egypt | 3.8 |
| Mauritania | 4.5 |
| UAE | 4.7 |
| USA | 4.7 |
| Europe | 5.4 |
| Bahrain | 6.4 |
| Oman | 6.7 |
| **Worldwide** | 15.5 |
| Sudan | 22.1 |
| Somalia | 35.6 |

| 70-85+ | |
|---|---|
| Libya | 2.6 |
| Palestine | 3.8 |
| Algeria | 4.5 |
| Tunisia | 4.6 |
| Lebanon | 5.3 |
| Syria | 5.4 |
| Jordan | 5.5 |
| Iraq | 6.0 |
| Bahrain | 7.4 |
| Mauritania | 7.7 |
| Morocco | 8.5 |
| USA | 10.7 |
| Europe | 11.2 |
| Egypt | 11.7 |
| S.Arabia | 12.6 |
| UAE | 15.5 |
| Oman | 15.9 |
| Kuwait | 23.4 |
| **Worldwide** | 29.0 |
| Sudan | 41.8 |
| Somalia | 44.0 |
| Qatar | 61.3 |

#### Males

| 20-39 | |
|---|---|
| Qatar | |
| Bahrain | |
| UAE | |
| Kuwait | 0.08 |
| Oman | 0.11 |
| Jordan | 0.13 |
| S.Arabia | 0.14 |
| Iraq | 0.15 |
| Libya | 0.15 |
| Palestine | 0.15 |
| Syria | 0.15 |
| Morocco | 0.16 |
| Egypt | 0.19 |
| Lebanon | 0.19 |
| Tunisia | 0.19 |
| Europe | 0.21 |
| Algeria | 0.23 |
| USA | 0.25 |
| Mauritania | 0.29 |
| **Worldwide** | 0.48 |
| Sudan | 0.76 |
| Somalia | 0.85 |

| 40-54 | |
|---|---|
| Bahrain | |
| UAE | 0.33 |
| Iraq | 0.50 |
| Qatar | 0.51 |
| Libya | 0.56 |
| Palestine | 0.67 |
| Kuwait | 0.69 |
| Algeria | 0.71 |
| S.Arabia | 0.80 |
| Tunisia | 0.92 |
| Syria | 0.97 |
| Jordan | 1.10 |
| Morocco | 1.10 |
| Egypt | 1.20 |
| Lebanon | 1.20 |
| Oman | 1.60 |
| Mauritania | 1.90 |
| USA | 5.40 |
| Europe | 6.50 |
| Sudan | 7.40 |
| **Worldwide** | 9.70 |
| Somalia | 11.70 |

| 55-69 | |
|---|---|
| Bahrain | 2.6 |
| Lebanon | 2.6 |
| Algeria | 3.0 |
| Qatar | 3.3 |
| Tunisia | 3.5 |
| Syria | 3.9 |
| S.Arabia | 4.0 |
| Oman | 4.0 |
| UAE | 4.1 |
| Iraq | 4.2 |
| Kuwait | 4.4 |
| Mauritania | 4.9 |
| Palestine | 5.0 |
| Jordan | 5.1 |
| Morocco | 6.1 |
| Egypt | 6.6 |
| Libya | 6.7 |
| Sudan | 14.2 |
| USA | 25.7 |
| Europe | 28.9 |
| Somalia | 32.4 |
| **Worldwide** | 42.4 |

| 70-85+ | |
|---|---|
| Palestine | 4.3 |
| Mauritania | 5.3 |
| Oman | 6.1 |
| Lebanon | 8.0 |
| Algeria | 8.0 |
| Syria | 8.7 |
| Tunisia | 8.8 |
| Iraq | 9.3 |
| Jordan | 11.7 |
| Morocco | 13.2 |
| Libya | 14.3 |
| Egypt | 15.1 |
| S.Arabia | 18.2 |
| Qatar | 22.1 |
| UAE | 23.9 |
| Bahrain | 25.5 |
| Kuwait | 30.5 |
| Sudan | 36.7 |
| Europe | 41.8 |
| USA | 44.7 |
| Somalia | 49.4 |
| **Worldwide** | 71.7 |

#### Oropharynx

##### Females

| 30-54 | |
|---|---|
| Qatar | |
| Palestine | |
| Jordan | |
| Libya | |
| Tunisia | |
| Kuwait | |
| Bahrain | |
| Mauritania | |
| Algeria | 0.02 |
| UAE | 0.06 |
| S.Arabia | 0.07 |
| Syria | 0.09 |
| Iraq | 0.10 |
| Egypt | 0.11 |
| Morocco | 0.13 |
| Somalia | 0.14 |
| Lebanon | 0.24 |
| Oman | 0.30 |
| Sudan | 0.32 |
| **Worldwide** | 0.41 |
| USA | 0.92 |
| Europe | 1.10 |

| 55-69 | |
|---|---|
| Qatar | |
| Palestine | |
| Kuwait | |
| Bahrain | |
| Mauritania | |
| UAE | |
| S.Arabia | 0.09 |
| Sudan | 0.26 |
| Tunisia | 0.28 |
| Somalia | 0.30 |
| Algeria | 0.31 |
| Jordan | 0.37 |
| Iraq | 0.42 |
| Egypt | 0.52 |
| Syria | 0.57 |
| Morocco | 0.60 |
| Lebanon | 0.65 |
| Oman | 0.98 |
| Libya | 1.40 |
| **Worldwide** | 1.70 |
| USA | 3.40 |
| Europe | 4.40 |

| 70-85+ | |
|---|---|
| Qatar | |
| Palestine | |
| Kuwait | |
| Bahrain | |
| Mauritania | |
| UAE | |
| Oman | |
| Libya | |
| S.Arabia | 0.38 |
| Egypt | 0.39 |
| Tunisia | 0.41 |
| Algeria | 0.55 |
| Jordan | 0.62 |
| Iraq | 0.66 |
| Syria | 0.75 |
| Morocco | 0.77 |
| Lebanon | 1.50 |
| **Worldwide** | 2.10 |
| Somalia | 2.20 |
| Europe | 3.20 |
| USA | 3.40 |
| Sudan | 4.00 |

##### Males

| 30-54 | |
|---|---|
| Qatar | |
| Palestine | |
| Bahrain | |
| Libya | |
| S.Arabia | 0.04 |
| UAE | 0.07 |
| Jordan | 0.09 |
| Kuwait | 0.10 |
| Egypt | 0.16 |
| Morocco | 0.19 |
| Sudan | 0.19 |
| Mauritania | 0.23 |
| Algeria | 0.25 |
| Somalia | 0.27 |
| Iraq | 0.28 |
| Syria | 0.31 |
| Tunisia | 0.33 |
| Lebanon | 0.59 |
| Oman | 0.61 |
| **Worldwide** | 1.80 |
| Europe | 4.30 |
| USA | 5.00 |

| 55-69 | |
|---|---|
| UAE | |
| Sudan | 0.22 |
| S.Arabia | 0.33 |
| Kuwait | 0.35 |
| Qatar | 0.55 |
| Egypt | 0.60 |
| Mauritania | 0.63 |
| Jordan | 0.67 |
| Palestine | 0.94 |
| Bahrain | 1.10 |
| Algeria | 1.10 |
| Somalia | 1.30 |
| Iraq | 1.30 |
| Syria | 1.30 |
| Libya | 1.60 |
| Tunisia | 1.60 |
| Oman | 1.60 |
| Lebanon | 2.00 |
| Morocco | 2.10 |
| **Worldwide** | 8.00 |
| Europe | 17.70 |
| USA | 19.10 |

| 70-85+ | |
|---|---|
| Qatar | |
| Mauritania | |
| Palestine | |
| UAE | |
| Sudan | |
| Kuwait | |
| Bahrain | |
| Libya | |
| S.Arabia | 0.85 |
| Somalia | 0.85 |
| Iraq | 1.40 |
| Egypt | 1.50 |
| Syria | 1.50 |
| Jordan | 1.70 |
| Oman | 2.00 |
| Lebanon | 2.30 |
| Tunisia | 2.90 |
| Algeria | 3.70 |
| Morocco | 3.90 |
| **Worldwide** | 8.30 |
| Europe | 10.90 |
| USA | 12.80 |

### Ovary

| 0-24 | | 25-54 | | 55-69 | | 70-85+ | |
|---|---|---|---|---|---|---|---|
| Tunisia | 0.22 | Oman | 3.3 | Oman | 11.2 | Iraq | 8.5 |
| Mauritania | 0.25 | S.Arabia | 3.4 | Tunisia | 12.6 | Mauritania | 11.5 |
| Kuwait | 0.31 | Kuwait | 3.7 | Mauritania | 12.6 | Algeria | 12.4 |
| Morocco | 0.31 | Tunisia | 4.2 | Iraq | 12.7 | Oman | 15.2 |
| Lebanon | 0.32 | UAE | 4.2 | Libya | 13.2 | Morocco | 16.1 |
| Syria | 0.37 | Qatar | 4.4 | S.Arabia | 13.5 | Tunisia | 16.9 |
| Jordan | 0.38 | [illegible] | 4.8 | [illegible] | 16.1 | S.Arabia | 18.1 |
| S.Arabia | 0.38 | Mauritania | 4.9 | Algeria | 16.6 | Jordan | 23.6 |
| UAE | 0.38 | Iraq | 5.4 | Kuwait | 17.8 | Syria | 23.6 |
| Oman | 0.39 | Libya | 5.5 | Egypt | 19.9 | **Worldwide** | 26.8 |
| Palestine | 0.40 | Jordan | 5.6 | Jordan | 20.9 | Libya | 27.5 |
| Algeria | 0.43 | Bahrain | 5.9 | Syria | 21.5 | Kuwait | 28.1 |
| Egypt | 0.43 | Algeria | 6.4 | **Worldwide** | 21.6 | Egypt | 28.7 |
| Sudan | 0.43 | Morocco | 6.5 | Morocco | 22.0 | Somalia | 28.8 |
| Iraq | 0.44 | Egypt | 7.0 | Qatar | 23.3 | Sudan | 29.4 |
| Libya | 0.46 | Syria | 7.3 | Somalia | 25.1 | [illegible] | 29.9 |
| Somalia | 0.46 | Sudan | 7.5 | Sudan | 25.5 | UAE | 29.9 |
| **Worldwide** | 0.48 | **Worldwide** | 7.8 | USA | 31.2 | Lebanon | 37.7 |
| USA | 0.48 | USA | 8.1 | Lebanon | 31.2 | Qatar | 38.8 |
| Europe | 0.54 | Somalia | 9.3 | UAE | 32.9 | Europe | 40.1 |
| Qatar | 0.77 | Lebanon | 10.5 | Europe | 33.7 | USA | 45.9 |
| Bahrain | 0.84 | Europe | 10.8 | Bahrain | 34.4 | Bahrain | 49.5 |

### Pancreas

#### Females

| 35-49 | |
|---|---|
| Qatar | |
| UAE | |
| Oman | 0.49 |
| Tunisia | 0.85 |
| S.Arabia | 0.86 |
| Mauritania | 0.90 |
| Bahrain | 0.97 |
| Palestine | 0.98 |
| Kuwait | 1.00 |
| Algeria | 1.10 |
| **Worldwide** | 1.10 |
| Iraq | 1.10 |
| Lebanon | 1.20 |
| Morocco | 1.20 |
| Sudan | 1.30 |
| Syria | 1.30 |
| Egypt | 1.40 |
| Jordan | 1.50 |
| Somalia | 1.50 |
| Libya | 1.70 |
| Europe | 2.20 |
| USA | 2.50 |

| 50-69 | |
|---|---|
| Sudan | 3.1 |
| S.Arabia | 5.3 |
| Mauritania | 5.4 |
| Morocco | 5.4 |
| Somalia | 5.8 |
| Tunisia | 6.1 |
| Iraq | 6.9 |
| Algeria | 7.1 |
| Qatar | 8.0 |
| Palestine | 8.6 |
| Libya | 8.6 |
| Syria | 9.1 |
| Egypt | 9.2 |
| Oman | 9.6 |
| Jordan | 9.6 |
| Lebanon | 10.7 |
| **Worldwide** | 11.5 |
| Kuwait | 13.1 |
| Bahrain | 13.8 |
| Europe | 20.2 |
| USA | 21.0 |
| UAE | 34.7 |

| 70-85+ | |
|---|---|
| Mauritania | 5.6 |
| Sudan | 7.5 |
| Somalia | 9.7 |
| Algeria | 13.5 |
| Morocco | 14.2 |
| S.Arabia | 14.9 |
| Jordan | 15.2 |
| Oman | 15.9 |
| Tunisia | 16.5 |
| Iraq | 18.2 |
| Syria | 18.5 |
| Bahrain | 18.8 |
| Egypt | 21.3 |
| Lebanon | 21.4 |
| Palestine | 40.6 |
| Libya | 41.7 |
| **Worldwide** | 48.4 |
| Kuwait | 50.8 |
| Qatar | 61.3 |
| Europe | 65.0 |
| USA | 67.7 |
| UAE | 114.4 |

#### Males

| 35-49 | |
|---|---|
| UAE | |
| Kuwait | 0.55 |
| Qatar | 0.61 |
| Bahrain | 0.76 |
| Iraq | 1.00 |
| Sudan | 1.00 |
| Somalia | 1.20 |
| Algeria | 1.40 |
| Morocco | 1.40 |
| Mauritania | 1.50 |
| S.Arabia | 1.50 |
| Oman | 1.80 |
| **Worldwide** | 2.10 |
| Tunisia | 2.50 |
| Palestine | 2.80 |
| Syria | 3.00 |
| Egypt | 3.30 |
| Jordan | 3.30 |
| Libya | 3.40 |
| USA | 3.50 |
| Europe | 3.90 |
| Lebanon | 4.60 |

| 50-69 | |
|---|---|
| Sudan | 4.0 |
| Somalia | 5.1 |
| Qatar | 6.2 |
| Mauritania | 6.9 |
| UAE | 7.6 |
| S.Arabia | 9.1 |
| Kuwait | 9.3 |
| Algeria | 10.1 |
| Iraq | 11.4 |
| Morocco | 11.4 |
| Oman | 12.8 |
| Tunisia | 12.8 |
| Bahrain | 13.5 |
| Palestine | 14.1 |
| Syria | 14.3 |
| Lebanon | 14.9 |
| Jordan | 16.2 |
| Libya | 16.4 |
| Egypt | 17.5 |
| **Worldwide** | 17.8 |
| USA | 30.7 |
| Europe | 32.7 |

| 70-85+ | |
|---|---|
| Sudan | 4.8 |
| Somalia | 11.1 |
| Mauritania | 11.1 |
| Qatar | 17.2 |
| Morocco | 17.9 |
| Tunisia | 18.1 |
| UAE | 19.6 |
| S.Arabia | 22.6 |
| Iraq | 23.8 |
| Algeria | 24.8 |
| Oman | 28.6 |
| Lebanon | 32.0 |
| Syria | 33.7 |
| Egypt | 34.3 |
| Palestine | 42.3 |
| Bahrain | 44.5 |
| Jordan | 45.9 |
| Kuwait | 46.5 |
| Libya | 53.2 |
| **Worldwide** | 57.0 |
| Europe | 83.8 |
| USA | 84.0 |

### Prostate

| 25-39 | | 40-54 | | 55-69 | | 70-85+ | |
|---|---|---|---|---|---|---|---|
| Qatar | | UAE | 0.65 | S.Arabia | 17.4 | Iraq | 80.5 |
| Palestine | | S.Arabia | 0.71 | Egypt | 26.8 | S.Arabia | 100.8 |
| Kuwait | | Kuwait | 0.81 | Iraq | 28.2 | Oman | 126.6 |
| Bahrain | | Egypt | 1.00 | Sudan | 32.5 | Sudan | 131.1 |
| Jordan | | Somalia | 1.30 | Bahrain | 34.8 | Egypt | 158.5 |
| Libya | | Qatar | 1.60 | Tunisia | 39.5 | Bahrain | 165.8 |
| Mauritania | | Iraq | 1.70 | Libya | 44.7 | Tunisia | 183.2 |
| UAE | | Bahrain | 1.80 | Algeria | 45.1 | Somalia | 190.6 |
| S.Arabia | 0.01 | Algeria | 1.80 | Somalia | 47.0 | Algeria | 191.6 |
| Morocco | 0.02 | Sudan | 1.90 | Jordan | 50.6 | UAE | 211.2 |
| Egypt | 0.04 | Jordan | 2.30 | Qatar | 53.6 | Jordan | 219.9 |
| Iraq | 0.05 | Libya | 2.70 | Oman | 61.2 | Qatar | 234.0 |
| Europe | 0.08 | Morocco | 2.80 | Palestine | 62.3 | Libya | 256.4 |
| Sudan | 0.08 | Palestine | 3.00 | UAE | 65.4 | Palestine | 268.8 |
| **Worldwide** | 0.11 | Tunisia | 3.00 | Kuwait | 71.1 | Syria | 272.4 |
| Oman | 0.15 | Mauritania | 3.10 | Mauritania | 74.4 | Morocco | 308.4 |
| Syria | 0.22 | Oman | 4.70 | Syria | 75.3 | Mauritania | 328.6 |
| Algeria | 0.23 | Syria | 5.40 | Morocco | 89.9 | **Worldwide** | 329.3 |
| Somalia | 0.23 | **Worldwide** | 8.90 | **Worldwide** | 132.8 | Kuwait | 341.5 |
| USA | 0.32 | Lebanon | 14.70 | Lebanon | 144.3 | Lebanon | 519.6 |
| Lebanon | 0.54 | Europe | 22.00 | Europe | 314.0 | USA | 534.0 |
| Tunisia | 0.69 | USA | 53.30 | USA | 410.5 | Europe | 595.8 |

### Salivary Glands

#### Females

### 30-44

| | |
|---|---|
| Qatar | |
| Bahrain | |
| UAE | |
| Morocco | 0.05 |
| Kuwait | 0.17 |
| Palestine | 0.24 |
| Sudan | 0.27 |
| Mauritania | 0.30 |
| Tunisia | 0.30 |
| Jordan | 0.32 |
| S.Arabia | 0.34 |
| Algeria | 0.40 |
| Egypt | 0.40 |
| **Worldwide** | 0.41 |
| Syria | 0.43 |
| Europe | 0.45 |
| Iraq | 0.45 |
| Oman | 0.56 |
| Somalia | 0.56 |
| Libya | 0.63 |
| USA | 0.66 |
| Lebanon | 0.69 |

### 45-59

| | |
|---|---|
| Qatar | |
| Bahrain | |
| Kuwait | 0.26 |
| Tunisia | 0.30 |
| UAE | 0.38 |
| S.Arabia | 0.48 |
| Algeria | 0.50 |
| Somalia | 0.51 |
| Sudan | 0.54 |
| Morocco | 0.61 |
| Iraq | 0.62 |
| Mauritania | 0.87 |
| Lebanon | 0.90 |
| **Worldwide** | 0.92 |
| Europe | 0.94 |
| Egypt | 0.99 |
| Syria | 1.00 |
| Palestine | 1.30 |
| Oman | 1.30 |
| USA | 1.30 |
| Libya | 1.50 |
| Jordan | 1.60 |

### 60-85+

| | |
|---|---|
| Qatar | |
| Bahrain | |
| Oman | |
| Libya | |
| Sudan | 0.50 |
| Algeria | 0.61 |
| Tunisia | 0.77 |
| Mauritania | 0.83 |
| Kuwait | 0.94 |
| S.Arabia | 1.10 |
| Iraq | 1.10 |
| Egypt | 1.10 |
| Jordan | 1.10 |
| Syria | 1.20 |
| Somalia | 1.50 |
| Lebanon | 1.50 |
| Morocco | 1.70 |
| Palestine | 1.90 |
| **Worldwide** | 2.10 |
| Europe | 2.10 |
| UAE | 2.60 |
| USA | 2.70 |

#### Males

### 30-44

| | |
|---|---|
| Libya | |
| Mauritania | |
| UAE | |
| Bahrain | |
| Kuwait | 0.12 |
| Qatar | 0.14 |
| Iraq | 0.16 |
| Tunisia | 0.16 |
| Algeria | 0.22 |
| Egypt | 0.22 |
| Sudan | 0.22 |
| S.Arabia | 0.23 |
| Somalia | 0.24 |
| Oman | 0.26 |
| Syria | 0.27 |
| Morocco | 0.30 |
| Lebanon | 0.31 |
| Jordan | 0.32 |
| **Worldwide** | 0.41 |
| Europe | 0.42 |
| Palestine | 0.51 |
| USA | 0.59 |

### 45-59

| | |
|---|---|
| Qatar | |
| Bahrain | |
| UAE | 0.06 |
| S.Arabia | 0.18 |
| Sudan | 0.20 |
| Egypt | 0.39 |
| Kuwait | 0.43 |
| Morocco | 0.50 |
| Libya | 0.54 |
| Oman | 0.54 |
| Algeria | 0.61 |
| Tunisia | 0.66 |
| Palestine | 0.88 |
| Mauritania | 0.89 |
| Iraq | 0.94 |
| Somalia | 1.00 |
| Jordan | 1.00 |
| **Worldwide** | 1.20 |
| Syria | 1.30 |
| USA | 1.50 |
| Europe | 1.60 |
| Lebanon | 1.70 |

### 60-85+

| | |
|---|---|
| Qatar | |
| Bahrain | |
| S.Arabia | 0.65 |
| UAE | 0.81 |
| Libya | 1.00 |
| Kuwait | 1.20 |
| Tunisia | 1.30 |
| Oman | 1.50 |
| Mauritania | 1.80 |
| Morocco | 2.00 |
| Jordan | 2.00 |
| Algeria | 2.10 |
| Iraq | 2.40 |
| Syria | 2.70 |
| Somalia | 2.80 |
| Egypt | 3.20 |
| Palestine | 3.40 |
| **Worldwide** | 3.40 |
| Lebanon | 3.50 |
| Sudan | 4.10 |
| Europe | 4.30 |
| USA | 5.80 |

#### Stomach

**Females**

| 20-39 | | 40-54 | | 55-69 | | 70-85+ | |
|---|---|---|---|---|---|---|---|
| Libya | 0.08 | S.Arabia | 1.4 | S.Arabia | 3.9 | Sudan | 5.8 |
| Sudan | 0.29 | Sudan | 1.8 | Sudan | 6.5 | Bahrain | 6.5 |
| S.Arabia | 0.32 | UAE | 2.6 | Kuwait | 7.0 | S.Arabia | 10.0 |
| Somalia | 0.38 | Qatar | 2.6 | Egypt | 8.2 | Morocco | 15.6 |
| Mauritania | 0.46 | Kuwait | 3.0 | USA | 9.5 | Egypt | 17.5 |
| UAE | 0.49 | USA | 3.4 | Bahrain | 9.5 | Iraq | 18.2 |
| Kuwait | 0.51 | Iraq | 4.1 | Iraq | 9.7 | Libya | 19.2 |
| Qatar | 0.52 | Tunisia | 4.1 | Tunisia | 10.7 | Palestine | 19.8 |
| USA | 0.70 | Somalia | 4.4 | Jordan | 11.7 | Kuwait | 20.3 |
| Oman | 0.82 | Libya | 4.5 | Morocco | 12.4 | USA | 24.5 |
| Egypt | 0.91 | Egypt | 4.5 | Libya | 12.5 | Jordan | 25.1 |
| Europe | 0.92 | Bahrain | 4.6 | Palestine | 13.2 | Algeria | 25.2 |
| Morocco | 0.92 | Mauritania | 5.9 | Syria | 14.4 | Somalia | 27.0 |
| Iraq | 0.96 | Europe | 5.9 | Algeria | 17.4 | Mauritania | 27.3 |
| Palestine | 1.00 | Morocco | 5.9 | Qatar | 19.0 | Tunisia | 27.4 |
| **Worldwide** | 1.10 | Palestine | 6.3 | Somalia | 19.2 | Qatar | 28.0 |
| Algeria | 1.20 | Jordan | 6.3 | Mauritania | 20.1 | Syria | 30.6 |
| Jordan | 1.30 | Oman | 6.4 | Europe | 20.7 | Oman | 42.8 |
| Tunisia | 1.40 | Algeria | 6.4 | Lebanon | 21.4 | Europe | 46.7 |
| Syria | 1.50 | Syria | 7.4 | UAE | 21.8 | Lebanon | 47.7 |
| Bahrain | 1.60 | **Worldwide** | 8.0 | Oman | 23.7 | **Worldwide** | 60.8 |
| Lebanon | 2.20 | Lebanon | 12.0 | **Worldwide** | 26.0 | UAE | 63.2 |

**Males**

| 20-39 | | 40-54 | | 55-69 | | 70-85+ | |
|---|---|---|---|---|---|---|---|
| UAE | 0.11 | UAE | 0.32 | S.Arabia | 9.6 | Bahrain | 17.6 |
| Bahrain | 0.13 | Bahrain | 1.10 | Sudan | 10.6 | Egypt | 22.6 |
| Qatar | 0.15 | Kuwait | 1.70 | Egypt | 10.8 | Sudan | 26.7 |
| Kuwait | 0.16 | Qatar | 1.80 | Kuwait | 10.9 | S.Arabia | 27.3 |
| Sudan | 0.31 | S.Arabia | 2.20 | UAE | 13.9 | Morocco | 31.1 |
| S.Arabia | 0.32 | Sudan | 4.20 | Qatar | 14.3 | Kuwait | 35.9 |
| Somalia | 0.47 | Somalia | 4.40 | Iraq | 18.2 | Iraq | 37.8 |
| Jordan | 0.50 | Egypt | 4.40 | Tunisia | 19.0 | Somalia | 41.1 |
| Egypt | 0.63 | Libya | 4.60 | Libya | 19.1 | Libya | 43.6 |
| Libya | 0.63 | Iraq | 5.30 | Somalia | 20.6 | Tunisia | 50.3 |
| Europe | 0.69 | USA | 5.70 | USA | 22.0 | Palestine | 50.8 |
| Iraq | 0.69 | Oman | 6.80 | Palestine | 22.3 | USA | 51.6 |
| USA | 0.69 | Tunisia | 7.00 | Bahrain | 23.3 | Mauritania | 52.4 |
| Mauritania | 0.75 | Palestine | 7.40 | Lebanon | 24.4 | Algeria | 53.9 |
| Syria | 0.79 | Jordan | 7.70 | Syria | 26.6 | UAE | 62.2 |
| Morocco | 0.80 | Algeria | 8.00 | Mauritania | 28.5 | Syria | 63.1 |
| Palestine | 0.82 | Mauritania | 8.20 | Algeria | 29.7 | Jordan | 73.4 |
| Algeria | 0.92 | Syria | 8.20 | Morocco | 29.9 | Lebanon | 76.1 |
| **Worldwide** | 1.00 | Morocco | 8.20 | Jordan | 37.9 | Oman | 87.8 |
| Oman | 1.10 | Europe | 10.60 | Europe | 50.0 | Qatar | 89.8 |
| Tunisia | 1.10 | Lebanon | 11.70 | Oman | 53.4 | Europe | 98.6 |
| Lebanon | 1.60 | **Worldwide** | 14.50 | **Worldwide** | 66.0 | **Worldwide** | 143.2 |

#### Testis

| 10-24 | | 25-54 | | 55-69 | | 70-85+ | |
|---|---|---|---|---|---|---|---|
| Mauritania | | Mauritania | | Qatar | | Qatar | |
| Somalia | 0.09 | Somalia | 0.46 | Oman | | Oman | |
| Libya | 0.11 | UAE | 0.60 | UAE | | UAE | |
| Sudan | 0.17 | Bahrain | 0.74 | Palestine | | Palestine | |
| Oman | 0.21 | Qatar | 0.76 | Egypt | 0.39 | Mauritania | |
| Algeria | 0.28 | Egypt | 0.77 | S.Arabia | 0.40 | Somalia | |
| Tunisia | 0.28 | Tunisia | 0.81 | Tunisia | 0.42 | Libya | |
| Egypt | 0.31 | Oman | 0.94 | Algeria | 0.55 | Jordan | |
| Morocco | 0.34 | Sudan | 0.97 | Morocco | 0.76 | Bahrain | |
| Bahrain | 0.41 | Algeria | 0.97 | Mauritania | 0.97 | Morocco | 0.28 |
| Qatar | 0.42 | Libya | 1.10 | Somalia | 1.00 | S.Arabia | 0.59 |
| Iraq | 0.51 | Kuwait | 1.10 | Libya | 1.00 | Egypt | 0.64 |
| S.Arabia | 0.71 | Morocco | 1.30 | Iraq | 1.10 | USA | 0.88 |
| Kuwait | 0.96 | S.Arabia | 1.40 | Syria | 1.10 | Algeria | 0.94 |
| Palestine | 1.20 | Iraq | 1.70 | Kuwait | 1.30 | Iraq | 0.94 |
| Syria | 1.20 | **Worldwide** | 3.00 | **Worldwide** | 1.30 | Sudan | 0.98 |
| **Worldwide** | 1.30 | Jordan | 3.00 | Jordan | 1.30 | **Worldwide** | 1.40 |
| Jordan | 1.40 | Palestine | 3.30 | Sudan | 1.40 | Europe | 1.50 |
| Lebanon | 1.60 | Syria | 3.90 | Lebanon | 1.50 | Tunisia | 1.60 |
| UAE | 1.60 | Lebanon | 7.40 | USA | 2.10 | Syria | 1.80 |
| USA | 4.60 | USA | 9.30 | Bahrain | 2.80 | Kuwait | 2.70 |
| Europe | 5.50 | Europe | 11.80 | Europe | 2.90 | Lebanon | 2.70 |

### Thyroid

#### Females

| 05-19 | | 20-39 | | 40-54 | | 55-69 | | 70-85+ | |
|---|---|---|---|---|---|---|---|---|---|
| Mauritania | | Mauritania | 0.76 | Mauritania | 2.2 | Mauritania | 5.3 | Mauritania | 3.8 |
| Sudan | 0.14 | Sudan | 1.40 | Sudan | 4.7 | Tunisia | 6.6 | Bahrain | 7.4 |
| Somalia | 0.23 | Somalia | 1.90 | Iraq | 5.9 | Iraq | 7.0 | Iraq | 7.5 |
| Libya | 0.24 | Libya | 3.10 | Tunisia | 6.3 | Libya | 8.8 | Somalia | 8.6 |
| Iraq | 0.27 | Iraq | 3.10 | Bahrain | 6.5 | Bahrain | 9.5 | Oman | 8.7 |
| Kuwait | 0.29 | Egypt | 4.20 | Somalia | 6.9 | Oman | 9.8 | Sudan | 10.5 |
| Egypt | 0.41 | Tunisia | 4.30 | Egypt | 7.5 | Egypt | 9.9 | Tunisia | 11.0 |
| Algeria | 0.56 | Bahrain | 5.80 | Libya | 7.6 | Sudan | 11.4 | Qatar | 12.8 |
| Tunisia | 0.58 | Qatar | 6.80 | Qatar | 10.6 | Somalia | 12.6 | Egypt | 17.3 |
| Morocco | 0.60 | UAE | 6.90 | Oman | 14.8 | Palestine | 13.3 | Algeria | 17.3 |
| Qatar | 0.73 | Algeria | 7.40 | Palestine | 15.2 | Syria | 14.6 | Syria | 17.5 |
| Bahrain | 0.83 | Morocco | 8.00 | Syria | 15.4 | Qatar | 16.9 | Europe | 17.5 |
| Palestine | 0.84 | Palestine | 9.00 | Jordan | 16.5 | Lebanon | 17.3 | **Worldwide** | 18.5 |
| Lebanon | 0.85 | Syria | 9.20 | Algeria | 16.7 | Jordan | 18.5 | Lebanon | 20.0 |
| **Worldwide** | 0.89 | **Worldwide** | 9.40 | Morocco | 21.2 | Algeria | 21.0 | Libya | 20.2 |
| Syria | 0.95 | Jordan | 10.10 | UAE | 22.3 | S.Arabia | 23.6 | Jordan | 22.3 |
| Jordan | 1.10 | Oman | 11.10 | **Worldwide** | 22.5 | Morocco | 23.8 | Morocco | 22.7 |
| Oman | 1.20 | Europe | 12.10 | S.Arabia | 23.0 | **Worldwide** | 25.0 | S.Arabia | 23.8 |
| S.Arabia | 1.70 | Kuwait | 13.00 | Europe | 23.5 | Europe | 25.6 | Kuwait | 32.2 |
| Europe | 1.80 | Lebanon | 13.20 | Kuwait | 24.7 | Kuwait | 33.7 | Palestine | 32.4 |
| UAE | 1.80 | S.Arabia | 13.90 | Lebanon | 25.5 | USA | 44.5 | USA | 32.4 |
| USA | 4.10 | USA | 26.50 | USA | 44.5 | UAE | 49.7 | UAE | 46.8 |

#### Males

| 05-19 | | 20-39 | | 40-54 | | 55-69 | | 70-85+ | |
|---|---|---|---|---|---|---|---|---|---|
| Mauritania | | Mauritania | 1.1 | UAE | 0.84 | Libya | 1.1 | Qatar | |
| Bahrain | 0.29 | Bahrain | 1.1 | Mauritania | 0.99 | Tunisia | 1.8 | Mauritania | 2.5 |
| Sudan | 0.38 | UAE | 1.4 | Bahrain | 2.80 | Mauritania | 2.5 | Libya | 3.6 |
| Libya | 0.41 | Tunisia | 1.6 | Morocco | 3.00 | Bahrain | 2.6 | Oman | 4.0 |
| UAE | 0.56 | Sudan | 1.9 | Kuwait | 3.20 | Iraq | 3.7 | Morocco | 4.6 |
| Somalia | 0.65 | Egypt | 2.5 | Libya | 3.30 | Jordan | 3.7 | Tunisia | 5.0 |
| Iraq | 0.74 | Somalia | 2.7 | Algeria | 3.40 | Qatar | 4.4 | Jordan | 5.1 |
| Tunisia | 0.80 | Libya | 2.8 | S.Arabia | 3.80 | Kuwait | 4.6 | Egypt | 5.5 |
| Qatar | 0.83 | Morocco | 3.0 | Jordan | 3.90 | Palestine | 4.6 | Somalia | 5.8 |
| Morocco | 0.91 | Algeria | 3.0 | Qatar | 4.00 | Egypt | 4.9 | Iraq | 6.1 |
| Egypt | 0.92 | Kuwait | 3.2 | **Worldwide** | 4.60 | Somalia | 4.9 | Syria | 6.9 |
| Oman | 0.97 | Iraq | 3.3 | Tunisia | 4.80 | Oman | 5.0 | Algeria | 7.2 |
| Kuwait | 1.40 | Qatar | 3.4 | Syria | 4.90 | Sudan | 5.4 | Palestine | 7.3 |
| Algeria | 1.40 | Oman | 3.4 | Egypt | 5.00 | Syria | 5.6 | UAE | 8.1 |
| Jordan | 1.60 | Jordan | 3.6 | Iraq | 5.20 | Algeria | 5.9 | **Worldwide** | 9.1 |
| Syria | 1.90 | Syria | 4.9 | Somalia | 5.30 | Morocco | 6.3 | Europe | 9.1 |
| Palestine | 2.10 | Palestine | 5.2 | Palestine | 5.50 | **Worldwide** | 8.8 | Kuwait | 9.7 |
| **Worldwide** | 2.30 | **Worldwide** | 6.2 | Europe | 6.00 | Lebanon | 9.4 | Lebanon | 9.8 |
| S.Arabia | 2.80 | S.Arabia | 6.3 | Oman | 6.30 | Europe | 9.5 | S.Arabia | 13.2 |
| Europe | 2.80 | Europe | 6.6 | Lebanon | 7.20 | UAE | 10.3 | Sudan | 13.3 |
| Lebanon | 3.20 | Lebanon | 8.4 | Sudan | 8.90 | S.Arabia | 10.9 | USA | 17.0 |
| USA | 5.80 | USA | 13.3 | USA | 9.10 | USA | 17.6 | Bahrain | 17.6 |

### Vagina

| 20-39 | | 40-54 | | 55-69 | | 70-85+ | |
|---|---|---|---|---|---|---|---|
| Qatar | | Qatar | | Qatar | | Qatar | |
| Oman | | Kuwait | | Kuwait | | Kuwait | |
| Kuwait | | Bahrain | | Bahrain | | Bahrain | |
| Bahrain | | Libya | | UAE | | UAE | |
| S.Arabia | | S.Arabia | 0.04 | Palestine | | Oman | |
| UAE | | UAE | 0.13 | S.Arabia | 0.21 | Mauritania | |
| Egypt | | Iraq | 0.16 | Tunisia | 0.65 | Iraq | 0.30 |
| Jordan | | Tunisia | 0.18 | Jordan | 0.76 | S.Arabia | 0.38 |
| Libya | | Algeria | 0.18 | Algeria | 0.80 | Sudan | 0.45 |
| Palestine | | Egypt | 0.30 | Iraq | 1.00 | Egypt | 0.74 |
| Tunisia | | Palestine | 0.37 | Syria | 1.10 | Algeria | 1.20 |
| Algeria | 0.02 | Syria | 0.39 | Egypt | 1.20 | Syria | 1.20 |
| Iraq | 0.02 | Europe | 0.43 | Europe | 1.20 | Palestine | 1.50 |
| Europe | 0.07 | Morocco | 0.52 | Oman | 1.20 | Tunisia | 1.50 |
| Morocco | 0.07 | Jordan | 0.56 | **Worldwide** | 1.30 | Jordan | 1.70 |
| **Worldwide** | 0.09 | **Worldwide** | 0.57 | Sudan | 1.40 | Somalia | 1.70 |
| Syria | 0.09 | Oman | 0.58 | Morocco | 1.50 | Europe | 2.30 |
| USA | 0.09 | Lebanon | 0.62 | Libya | 1.60 | **Worldwide** | 2.40 |
| Sudan | 0.12 | USA | 0.67 | Lebanon | 1.60 | Morocco | 2.40 |
| Mauritania | 0.16 | Somalia | 0.67 | USA | 1.60 | Lebanon | 2.50 |
| Lebanon | 0.19 | Sudan | 0.77 | Somalia | 2.00 | Libya | 2.60 |
| Somalia | 0.28 | Mauritania | 1.10 | Mauritania | 2.20 | USA | 3.30 |

#### Vulva

| 20-39 | | 40-54 | | 55-69 | | 70-85+ | |
|---|---|---|---|---|---|---|---|
| Qatar | | Qatar | | Qatar | | Qatar | |
| Kuwait | | Libya | | S.Arabia | 0.34 | Kuwait | |
| Libya | | Bahrain | | UAE | 0.52 | Oman | |
| Bahrain | | UAE | | Iraq | 0.55 | UAE | |
| Egypt | | S.Arabia | 0.16 | Mauritania | 0.61 | Bahrain | |
| UAE | | Kuwait | 0.27 | Libya | 0.69 | Iraq | 0.98 |
| Iraq | 0.02 | Egypt | 0.29 | Palestine | 0.85 | S.Arabia | 1.50 |
| S.Arabia | 0.02 | Tunisia | 0.34 | Algeria | 0.86 | Jordan | 1.50 |
| Algeria | 0.05 | Palestine | 0.37 | Kuwait | 0.94 | Mauritania | 2.10 |
| Jordan | 0.07 | Algeria | 0.39 | Jordan | 1.10 | Algeria | 4.60 |
| Morocco | 0.07 | Jordan | 0.42 | Oman | 1.20 | Tunisia | 5.20 |
| Tunisia | 0.11 | Iraq | 0.48 | Bahrain | 1.50 | Somalia | 6.00 |
| Syria | 0.12 | Mauritania | 0.70 | Tunisia | 1.50 | Egypt | 6.50 |
| Oman | 0.15 | Syria | 0.74 | Egypt | 1.60 | Palestine | 6.80 |
| Palestine | 0.15 | Morocco | 0.93 | Syria | 2.40 | **Worldwide** | 7.60 |
| Sudan | 0.21 | Sudan | 0.97 | **Worldwide** | 2.80 | Sudan | 7.70 |
| Lebanon | 0.25 | **Worldwide** | 1.10 | Morocco | 3.70 | Syria | 7.80 |
| Mauritania | 0.30 | Lebanon | 1.30 | Somalia | 4.80 | Libya | 9.20 |
| **Worldwide** | 0.31 | Somalia | 1.30 | Europe | 5.50 | USA | 11.10 |
| Europe | 0.40 | Oman | 1.40 | Lebanon | 5.70 | Morocco | 12.50 |
| USA | 0.42 | Europe | 2.40 | USA | 6.00 | Europe | 15.10 |
| Somalia | 0.64 | USA | 2.80 | Sudan | 6.30 | Lebanon | 19.50 |
